# A signal-seeking Phase II trial of Durvalumab and Tremelimumab Focused on Advanced, Rare and Less Common Cancers

**DOI:** 10.1101/2022.06.30.22277092

**Authors:** Subotheni Thavaneswaran, Frank P Lin, Maya Kansara, John P Grady, David Espinoza, Anthony M Joshua, Peter Grimison, Paul Craft, Rasha Cosman, Chee Lee, Kathleen Harwood, Sarah Chinchen, Theresa Corpuz, Mandy Ballinger, Lucille Sebastian, John Simes, David Thomas

## Abstract

Immune checkpoint blockade impedes the negative regulatory signals for T-cell response and permits more effective immune detection and eradication of cancer cells. This single-arm phase II clinical trial (ACTRN12616001019493) within the Molecular Screening and Therapeutics (MoST) program evaluates the clinical activity and safety of combination immunotherapy with durvalumab and tremelimumab in patients with advanced cancers, prioritsing rare cancers (<6 per 100,000 annual incidence) and patients having failed standard treatments for their cancer type.

**Methods:** Eligible patients were determined by the molecular tumour board based on the absence of actionable genomic findings (n=64) and biomarker enriched (n=48) at screening. Patients received durvalumab 1500 mg and tremelimumab 75 mg every four weeks for 4 cycles, followed by durvalumab alone for another 9 cycles. The primary endpoint was progression-free survival at 6 months (PFS6) and secondary endpoints included objective response, time to progression (TTP) on trial to TTP on prior therapy (TTP2/TTP1>1.3), overall survival and treatment tolerability.

**Results:** Between December 2016 and 2019, 112 patients were enrolled on the study. There was a female predominance (55%), most had an ECOG performance status of 0 (66%), aged <65 years (75%), with rare cancers (84%). The PFS6 rate was 32% (95% CI 23 to 40%); 16 of 112(14%) achieved an objective response; TTP2/TTP1>1.3 for 22 of 63 (35%) patients with an evaluable ratio; median overall survival 11.9 months (95% CI 11.0 to 14.8), and there were no new safety concerns. High tumour cell PD-L1 correlated with improved PFS and OS and TMB with PFS alone. More PD-1^+^CD4^+^ T-cells and circulating follicular T-helper (cTfh) cells at baseline were strongly associated with better PFS and OS.

**Conclusion:** Durvalumab plus tremelimumab demonstrated a signal of clinical activity in treatment-refractory patients with rare cancers. A PFS6 of 32% and 35% of patients achieving a TTP2/TTP1>1.3 suggests an improved disease trajectory on trial. Translational correlates provided insights into biological associations with clinical outcomes across tumour types.

## INTRODUCTION

Checkpoint inhibitors target tumour immune evasion, which appears broadly relevant to cancer development (1). Durvalumab and tremelimumab are both humanised immunoglobulin G monoclonal antibodies that bind respectively to the programmed death-ligand 1 (PD-L1) and the coinhibitory receptor CTLA-4, which are suppressors of T-cell activity in cancers (2,3). Despite the targeted nature of these therapies, a clear biomarker has not been identified across cancer types. While high PD-L1 expression or high tumour mutational burden (TMB) may increase the likelihood of favourable clinical outcomes in select cancer histologies, their presence or absence does not guarantee, or preclude a response (4,5). Pembrolizumab (PD-1 inhibitor) monotherapy has received FDA tumour agnostic approval for both mismatch repair deficiency/microsatellite instability, and increased TMB (≥ 10 mutations/megabase) using the FoundationOne CDx™ assay. This approval was based on the KEYNOTE-158 study, which included a limited sampling of 27 cancer types, with half represented by 4 cancers (endometrium, pancreas, biliary, gastric) (6). Sarcomas, for example, comprise 50 histologic types and represented 10% of the cohort. Moreover, given histotypic variation in TMB and lack of standardization across sequencing platforms, the rationale for a TMB cut-off of 10 mutations/megabase across cancer types remains unclear (7-9). Finally, the activity of dual checkpoint blockade in a tumour agnostic setting, particulary in rare and less common cancers is unknown.

We have conducted a tumour agnostic, signal-seeking trial of durvalumab and tremelimumab in mostly treatment-refractory patients, prioritising rare and less common cancers due to their under-representation in clinical trials. This trial formed part of the Molecular Screening and Therapeutics study (10), in which patients underwent comprehensive genomic profiling (CGP) prior to enrolment onto a range of therapeutic substudies. The initial cohort of 64 subjects was unselected for biomarkers, while an expansion cohort of 48 subjects was enriched for intermediate/ high TMB. A range of correlative biomarkers were retrospectively examined for an association with clinical outcomes.

## METHODS

### Patients

Patients were treated at five sites across Australia. The study was performed in accordance with the Declaration of Helsinki. Central or institutional ethics and local research governance approval were obtained. All patients provided written informed consent for participation in this trial. An independent data and safety monitoring committee provided independent assessments of patient safety and trial progress. Key eligibility criteria included patients with pathologically confirmed, advanced or metastatic solid cancer of any histological type, excluding cancer types for which immunotherapy is already standard of care in Australia; Eastern Cooperative Oncology Group performance status 0–1; adequate hepatic, renal and marrow function. Patients were required to be progressing on, be intolerant of, or have documented unsuitability for further standard therapy for their tumour type and not have previously received treatment with a PD1, PD-L1, or CTLA-4 inhibitor. Documented unsuitability includes known hypersensitivity, organ dysfunction, or other patient factors that would make standard therapy unsuitable in the judgement of the responsible investigator.

### Screening status

CGP was undertaken as part of the molecular screening and therapeutics (MoST) program using formalin-fixed, paraffin-embedded archival tumour (10). The panel assay employed for MoST screening evolved over time and included three in-house assays (MoST, CCPv2, CCPv2.2), Illumina TruSight Tumor 170, and Foundation Medicine (FMI). Molecular eligibility was determined by the Molecular tumour board (MTB), with either no actionable mutations to enrol on a biomarker-specified MoST trial (initial cohort, n=64), or enriched for intermediate/high TMB (expansion cohort, n=48). Patients enrolled on trial were also required to have additional tissue available for retrospective biomarker assessment.

The TMB was initially estimated according to the whitepaper methods outlined by Illumina (11). In brief, all coding mutations were included and filtered to remove low variant allele frequency (VAF) variants (VAF ≥ 0.05) and population variants (gnomAD frequency ≥ 0.001%). However, variants with high Catalogue of Somatic Mutations in Cancer (COSMIC) count ≥ 20 were re-introduced to calculate TMB as the number of somatic mutations per Mb. This was later revised to remove driver mutations (COSMIC count > 1) in accordance with Lieber et al (12) and reduce the ascertainment bias of sequencing known cancer genes. Due to the small panel size of the TST170 (0.56Mb), we chose to retain synonymous mutations in the calculation, despite no expected contribution to neo-antigen production, to increase the sample size of somatic mutations per patient and reduce noise in the calculation. There was also post-hoc harmonisation across sequencing panels employed (13-15).

### Design and statistical considerations

This trial is a multi-centre, open-label, phase IIa trial comprised of 7 modules of 16 patients each. Of a total of 112 patients treated on trial, the first cohort comprised of 4 modules of 16 patients (n=64) with a post-hoc analysis planned to allocate patients into 4 subgroups based on tumour cell PD-L1 expression and TILs, using median expression levels. Interim analyses and external data led to an expansion cohort enriched for intermediate/high TMB, comprised of 3 modules of 16 patients (n=48) and a change in primary endpoint to progression-free survival at 6 months (PFS6). The rationale for this change was based on ORR having limited scope in capturing the clinical benefit of immunotherapy, but a threshold for clinical activity using PFS6 was not set.

### Treatment

All patients were treated with a fixed dose of both 1500 mg durvalumab intravenously (IV) and 75mg of tremelimumab IV (D+T) every 4 weeks for the first four cycles. After completion of 4 cycles, durvalumab alone was continued every 4 weeks for up to an additional 9 doses. Treatment continued until disease progression, unmanageable toxicity, a decision by the patient or clinician to stop, or completion of the 13 cycles of trial protocol therapy. Patients who achieved and maintained disease control through to the end of the 12-month treatment period were permitted to restart combination treatment with D+T upon evidence of disease progression, as were patients who demonstrated progressive disease whilst receiving durvalumab monotherapy.

### Endpoints

The primary trial endpoint was progression-free survival at 6 months (PFS6). Secondary objectives included objective tumour response, overall survival (OS), ratio of time to progression (TTP) on trial (TTP2), to TTP on the last line of therapy (TTP1) prior to trial entry; safety and tolerability of treatment; and health-related quality of life (QoL). QoL was defined by scores on the EORTC QLQ-C30, and Brief Pain Inventory (BPI) (16), assessed at baseline and every 4 weeks until disease progression. In a pan-cancer setting, using patients as their own control informs the rate of change in disease trajectory for that individual, with a TTP2/TTP1 ratio of 1.3 suggesting clinical activity (17,18).

Pre-specified analyses of biomarkers examined the potential predictive value of PD-L1 expression, TILs and TMB by comparing differences on progression-free and overall survival for these biomarkers. Additional exploratory analyses of biomarkers potentially predictive of response to immunotherapy were undertaken, including histotype, immune phenotyping and gene mutations including mismatch repair deficiency (MMRD) by sequencing. Examination of tumour cell PD-L1 expression was by immunohistochemistry using the Ventana PD-L1 (SP263) assay and a cut-off for positivity set at ≥1%, based on the distribution of PD-L1 across the study cohort. Hematoxylin and eosin-stained slides were used for morphological discrimination of lymphocytes in the tumour and its immediate periphery, as previously described (19,20) and the tumour-infiltrating lymphocytes (TILs) level was quantified as proportion TILs, of total cells on a slide, and dichotomised as low and high using median TILs for the cohort. TMB measures used to qualify patients for the expansion trial cohort were determined by the MTB. In post-hoc analysis, TMB was harmonized by first converting panel-specific values to whole-exome sequencing (WES) equivalent TMB values (14,15), before converting to Foundation Medicine equivalent TMB (muts/MB) where the cut-off for intermediate/high TMB was set at ≥10 mutations per megabase (21).

Lastly, an exploration of the immune phenotype of peripheral blood mononuclear cells (PBMCs) was undertaken using baseline and week 4 samples to identify early immune activation and associated biomarkers of benefit from dual immune checkpoint blockade. Bloods were collected in sodium-heparin tubes and PBMCs were stored frozen until analysed by flow cytometry. Samples were excluded from statistical analyses if the event count of the parent was <50. These biological parameters were then assessed according to the overall clinical outcome (Good = objective response, SD>12 months; Intermediate = SD, TTP> 3 months; and Poor = PD, TTP<3 months), OS and PFS. Details of flow cytometry methods and statistical analysis are described in Appendix 5.

## RESULTS

### Cohort characteristics

Patients were enrolled in two cohorts, the first between December 2016 and December 2017 and the second between November 2018 and November 2019. The median age was 54 years (range 18 to 84 years), 49 (44%) were male and 63(56%) had an ECOG PS of 0. The most common histotypes included bone and soft tissue sarcomas (50, 45%), primary central nervous system tumours (10, 9%), and colorectal adenocarcinomas (10, 9%). Patients were heavily pretreated, with a median of 3 lines of previous systemic therapy (range 0-15) (Table 1). CGP results were available for 105 patients, 55 sequenced on an in-house assay (46 on CCPv2/2.2 and 9 on MoST), 10 on FMI and 40 on TST170. At the time of MTB report issue, 44 patients were considered to have an intermediate/ high TMB (Appendix 1, Supplementary Table 1)

**Table 1.** Baseline characteristics (N=112)

| Characteristic | No. | % |  |
| --- | --- | --- | --- |
| Male sex | 49 | 44 |  |
| Median age (range) | 53.9<br>(39.8 – 64.7) |  |  |
| ECOG status |  |  |  |
| 0 | 74 | 66 |  |
| 1 | 38 | 34 |  |
| Median lines of prior systemic therapy (range) | 3 (0-15) |  |  |
| Cancer type |  |  | Classifications by incidence – rare: <6 per 100,000; less common 6-12 per 100,000; common >12 per 100,000. |
| Adenoid cystic carcinoma | 4 | 3.6 | Rare |
| Carcinoma of unknown primary | 5 | 4.5 | Rare |
| Biliary tract | 4 | 3.6 | Rare |
| Brain, glioblastoma | 6 | 5.4 | Rare |
| Brain, other gliomas | 3 | 2.7 | Rare |
| Colorectal adenocarcinoma | 10 | 8.9 | Common |
| Prostate adenocarcinomas | 4 | 3.6 | Common |
| Other adenocarcinomas | 7 | 6.3 | Rare |
| Cutaneous, squamous cell carcinoma (SCC) | 2 | 1.8 | Common |
| Other SCC | 2 | 1.8 | Rare |
| Esophagogastric | 2 | 1.8 | Less common |
| Head & Neck carcinoma | 2 | 1.8 | Rare |
| Other carcinomas | 6 | 5.4 | Rare |
| Ovarian | 3 | 2.7 | Rare |
| Primitive neuroectodermal tumour | 4 | 3.6 | Rare |
| Sarcoma, leiomyosarcoma | 13 | 11.6 | Rare |
| Sarcoma, undifferentiated | 7 | 6.3 | Rare |
| Sarcoma, WDLPS | 5 | 4.5 | Rare |
| Sarcoma, other | 21 | 18.7 | Rare |
| Other | 2 | 1.8 | Rare |

WDLPS – well-differentiated/ de-differentiated liposarcoma. Other comprised of a Wilms tumour and a pituitary adenoma.
Excluded cancer types: cutaneous melanoma, non-small cell lung cancer, squamous cell carcinoma of the head and neck, breast cancer, bladder cancer, thymoma, thymic carcinoma or pancreatic cancer.

### Primary and secondary endpoints

With a median follow-up time of 28 months, the primary endpoint of PFS rate at 6 months was 32% (95% confidence interval (CI), 23-40%). The median PFS was 2.92 months (95% CI 1.84-3.61 months; Figure 1). Of the 112 patients, 10 did not have measurable disease by RECIST/RANO criteria; or further imaging following trial enrolment. Nine patients achieved a partial response and seven patients a complete response, equating to an objective response rate of 14% (16 of 112). Objective responses were seen in rare histotypes including two epithelioid hemangioendotheliomas, and one each of angiomatoid fibrous histiocytoma, pleomorphic sarcoma, merkel cell carcinoma, pancreatic acinar carcinoma, and ampullary carcinoma (Figures 2 and 3). Forty patients (36%) achieved stable disease (SD) and 3 patients (3%) a non-complete response/non-progressive disease (non-CR/non-PD) as best response. The remaining 43 patients (42%) demonstrated progressive disease as best response; 37 by RECIST assessment and 6 by RANO (Figure 2). The 35 patients that remained progression-free at 6 months comprised 16 patients who achieved an objective response, 17 patients with SD, and two with non-CR/non-PD. The median OS was 11.9 months (95% CI 11.0 to 14.8 months) (Figure 1B).

**Figure 1A.**
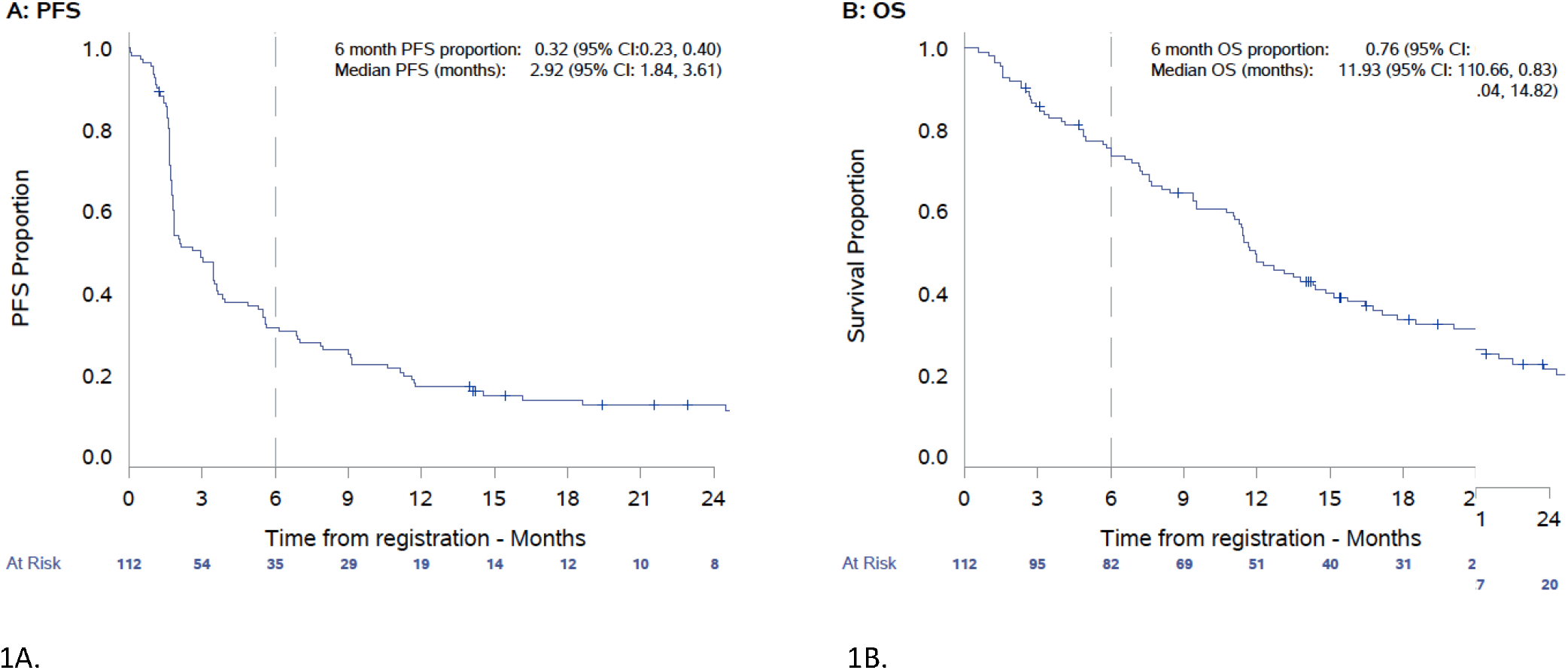
Progression-free survival (PFS), including primary endpoint of 6 month PFS. **1B**. Overall survival.

**Figure 2.**
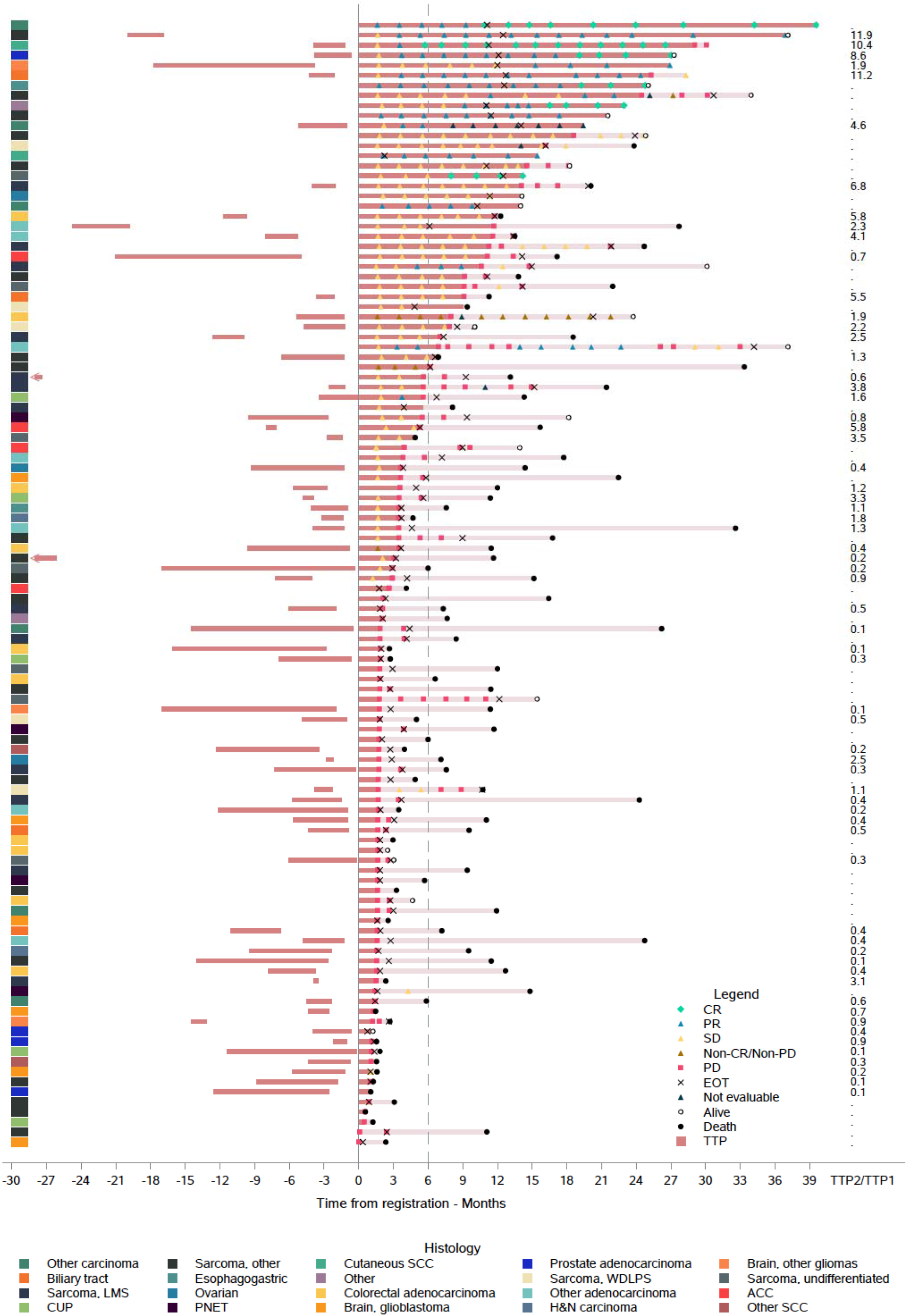
Swimmerplot including TTP1 and TTP2/1 ratio where evaluable, best response and duration of response by histotype. Months nominated by – indicate TTP1 time in months. Abbreviations: CR – complete response, PR – partial response, SD – stable disease, PD – progressive disease, EOT – end of treatment, TTP – time to progression, WDLPS – well-differentiated liposarcoma, PNET – primitive neuroectodermal tumor, SCC – squamous cell carcinoma, L leiomyosarcoma, ACC – adenoid cystic carcinoma, H&N – head and neck, CUP – carcinoma of unknown primary site.

**Figure 3.**
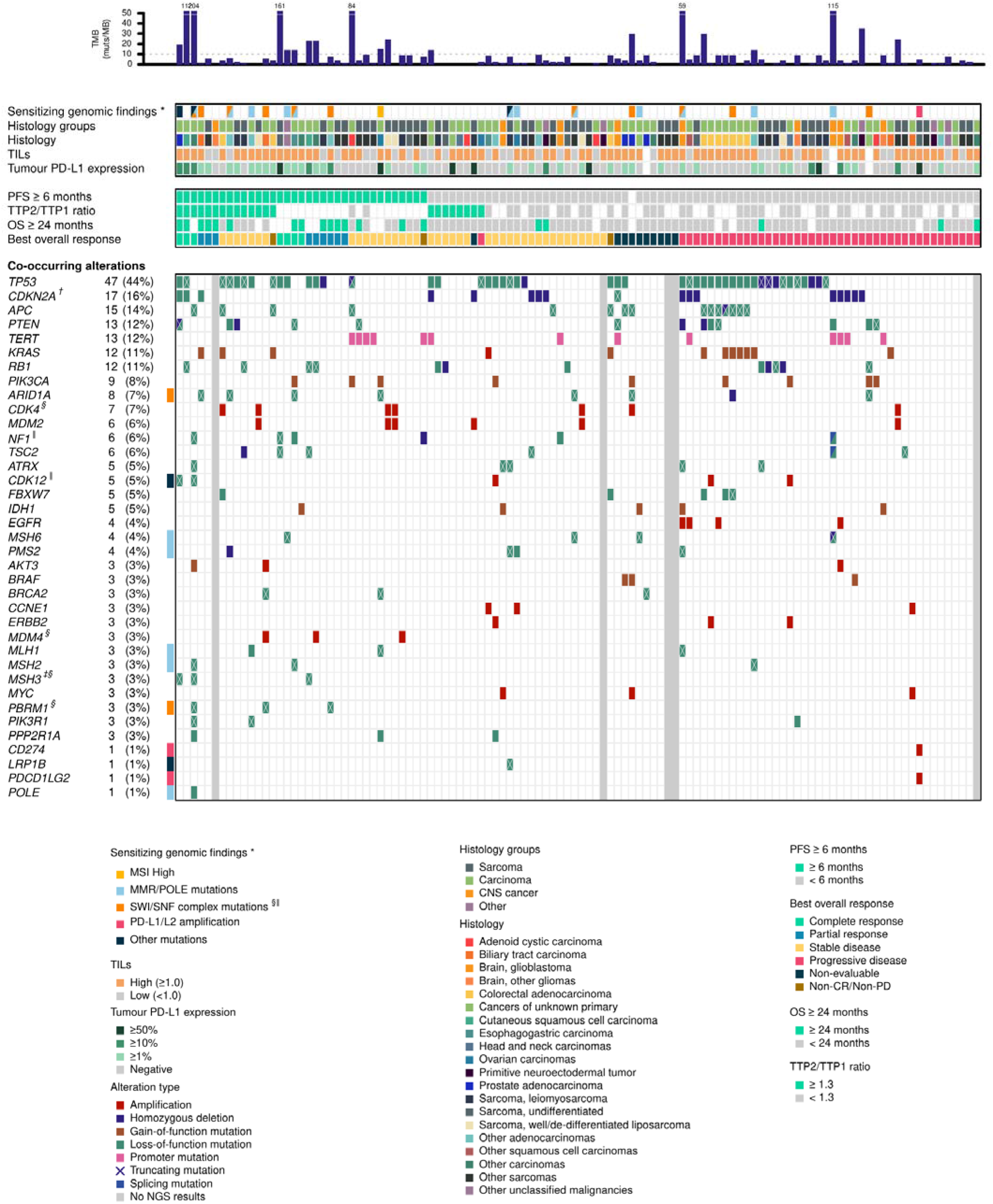
Heatmap of genomic alterations with immunopathologic assays and clinical outcomes. Only alterations present in ≥ 3 cases and genes of special interest are shown in the heatmap. Tumour mutational burden (TMB) were normalised to non-synonymous mutations per megabase; high TMB (orange bars) defined as ≥10 muts/Mb. Notes: (*) potentially sensitizing genomic findings include microsatellite instability-high (MSI-H), genomic alterations in the mismatch repair gene (MMR genes, including MLH1, MSH2, MSH6, PMS2) and/or POLE mutation, alteration in genes encoding the switch/sucrose non-fermentable (SWI/SNF) complex (ARID1A, PBRM1), PD-L1/L2 (CD274, PDCD1LG2) genomic amplifications, and others (CDK12, LRP1B). Fisher’s exact test was used to examine the association between genomic alterations and (†) PFS6 and (‡) best overall response (BOR) at type I error rate α of 0.01, respectively; (§) and (⍰) indicate a trend toward statistical significance (0.01≤α< 0.05) with PFS6 and BOR, respectively. Abbreviations: OS: overall survival; PFS: progression-free survival; TIL: Tumour infiltrating lymphocytes; TTP: time-to-progression; TTP2/TTP1: ratio of TTP of durvalumab and tremeliumab to TTP of the immediate preceeding systemic treatment.

To account for the heterogeneity of cancer types and natural histories, we estimated the TTP2/TTP1 ratio for 63 patients (Appendix 2). The median TTP1 for the cohort was 3.9 months (range 0.5-18.0 months). A TTP2/TTP1 ratio of >1.3 (the pre-defined threshold for clinical activity) was achieved in 22 patients (Figure 2). In the subset of patients achieving SD as the best overall response, the median TTP2/TTP1 ratio was 1.6 (range 0.17-6.8), with a median OS of 14.8 months (95% CI 12.3 to 18.5). Of interest, amongst the 35 patients who met the primary PFS6 endpoint, the median TTP2/1 ratio was 4.4 (range 0.7-11.9 months) with 14 patients of 16 with an evaluable ratio (88%) achieving a TTP ratio>1.3, indicating an improved disease trajectory on study.

### Safety and adverse events

All 112 patients (100%) experienced at least one adverse event (AE) of any grade while receiving trial therapy. Forty seven serious AEs were experienced by 30 patients (27%), with only 17 of these considered related to trial drugs. Immune-related serious AEs were considered of special interest and included colitis (n=3), hepatitis (n=3), pancreatitis (n=2), pneumonitis (n=3) and other auto-immune events (n=4) (Appendix 3, Supplementary Table 3). There were two grade 5 serious AEs – cardiac arrest and intracranial haemorrhage, which were both considered unrelated to study treatment.

### Patient reported outcomes

Global health status based on the QLQ-C30 demonstrated a mean decline of -4.2 (95% CI -8.10 to - 0.23) compared to baseline. Role functioning declined with a mean change of -6.5 (95% CI -11.1 to - 1.9) from baseline, while there was no evidence of a change in cognition (−0.76, 95% CI -3.8 to 2.3) or emotional functioning (+1.7, 95% CI -1.7 to 5.1). Importantly, no deterioration was evident in the Brief Pain Inventory mean pain severity, -0.06 (95% CI -0.62 to 0.73) or pain interference, -0.14 (95% CI -0.63 to 0.35). Mean global health status declined by -8.73 (95% CI: -13.63. to -3.83) amongst patients progressing before 6 months, while the 35 patients who remained progression-free at 6 months experienced a mean improvement in global health status of 4.81 (95% CI: -0.87 to 10.48). Overall, none of these changes from baseline in global health status or subscales met thresholds for clinically meaningful differences (22, 23).

### Exploratory endpoints

Tumour cell PD-L1 expression was evaluable in 109 (97%) patients. Median PD-L1 level was 0.5% (range 0 to 100%) and comprised of 54% with a PD-L1 expression <1% and 46% with a PD-L1 level ≥1%. Patients with tumour cell PD-L1 expression ≥1% had a longer median PFS of 5.5 (95% CI 1.8 to 1.0) months compared with 2.0 (95% CI 1.8 to 3.5) months in those with PD-L1 expression <1%, HR: 0.56 (95% CI 0.37-0.85, logrank P=0.0061). Similarly, median OS was significantly longer in patients with tumour cell PD-L1 ≥1% (18.5 months, 95% CI 11.2 to 27.7) compared to those with PD-L1 expression <1% (9.5 months, 95% CI 7.1 to 12.7), HR: 0.50, 95% CI 0.31-0.78, logrank P=0.0021) (Figure 4A and B). Furthermore, tumour cell PD-L1 expression was a significant predictor of both objective response (ROC AUC, 0.723, 95% C.I. 0.600-0.846) and PFS6 (AUC 0.624, 95% CI 0.518-0.730); Appendix 4, Supplementary Figure 1. At the threshold of ≥1%, the sensitivity and specificity were 0.87 and 0.59, respectively, for predicting the best overall response. Median TILs in 109 evaluable subjects was 1.0 (IQR 0.0 to 5.0). No association between TILs >1 and PFS, HR 0.83 (95% CI 0.55 to 1.26) or OS, HR 0.83 (95% CI 0.53 to 1.31) was seen (Figures 4C and D).

**Figure 4.**
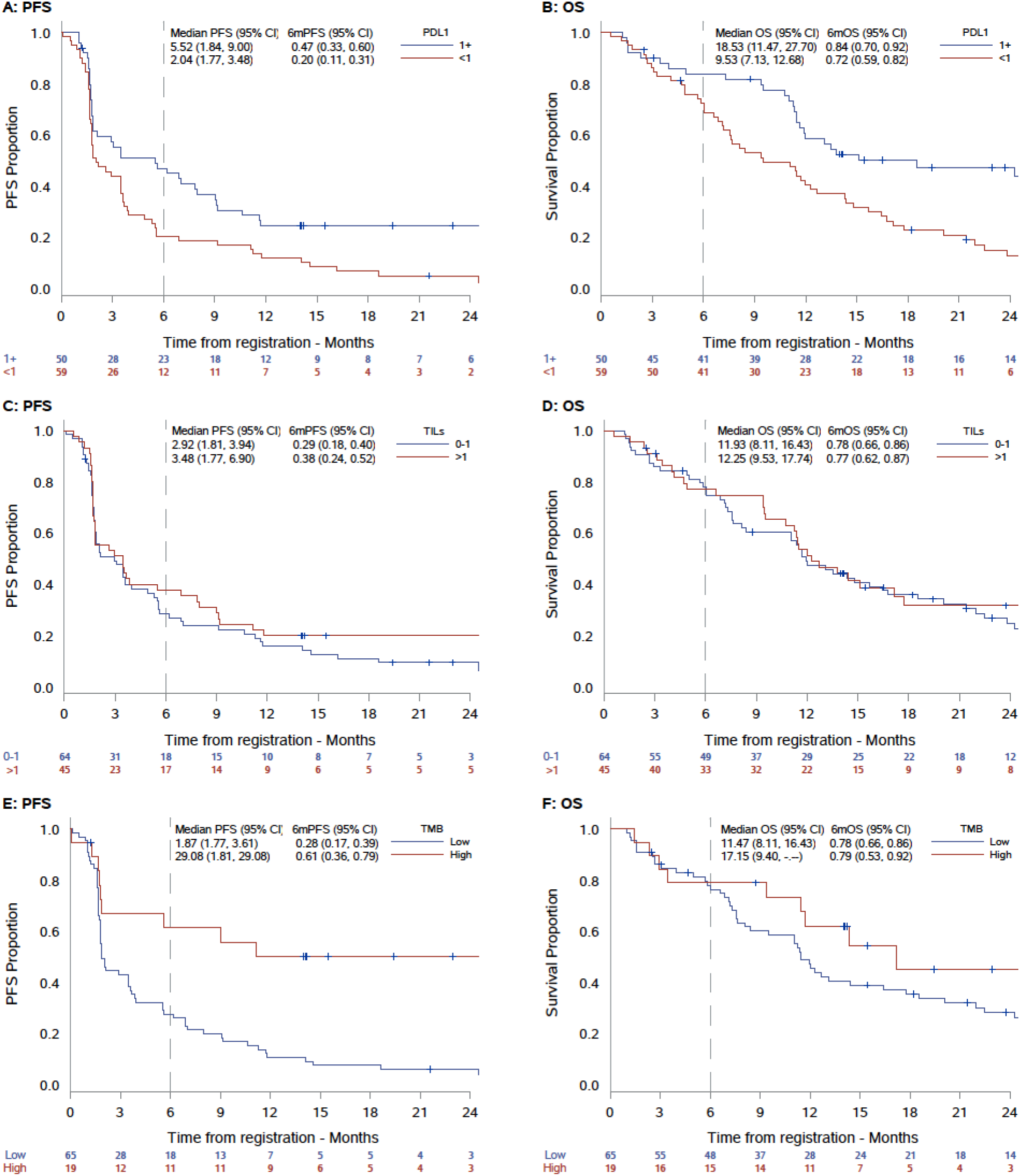
A and B. PFS and OS by PD-L1. 4C and D. PFS and OS by TILs. 4E and F. PFS and OS by TMB.

The results of CGP was available for 105 patients, and a TMB result (both prospectively and retrospectively calculated) was available for 82 patients (78%; 38 on CCPv2/v2.2, 10 on FMI and 34 on TST170). For the original 64-patient cohort, TMB was only computed retrospectively at the time of study analysis, with most patients sequenced on the CCP panel. For the expansion cohort, TMB was reported at time of MTB issue for the majority, but underwent subsequent harmonisation as detailed in Appendix 1. Sixteen of 39 patients in the expansion cohort with an intermediate/high TMB at time of MTB issue maintained an intermediate/high TMB following post-hoc harmonisation. Amongst the 19 patients with a harmonised TMB ≥10 mutations/Mb across assays, median PFS was 29.1 months, compared with 2.0 months for those with low TMB (P=0.0004, logrank test). There was also a trend towards improved OS of 17.2 months for tumours with high TMB compared with 11.5 months for low TMB tumours (logrank P=0.11) (Figure 4E and F). TMB was not significantly associated with either objective response (AUC 0.657, 95% CI: 0.465-0.874), or PFS6 (AUC 0.618, 95% CI: 0.484-0.752); Appendix 4, Supplementary Figure 2. When grouped by low (<10 mutations/Mb), intermediate (10-20 mutations/Mb) and high TMB (>20 mutations/ Mb), TMB was associated with objective response (P=0.0035) and a trend towards an improved PFS6 rate (P=0.055), although the intermediate TMB performed better than the high. In a multi-variable analysis of these exploratory biomarkers, only tumour cell PD-L1 expression ≥1% maintained a significant association with both PFS and OS.

Of note, mismatch repair deficiency correlated inconsistently with TMB in this cohort, with 15 individuals carrying mutations in mismatch repair genes (*MLH1, MSH2, MSH6, PMS2 or POLE*). Deletions (but not point mutations) in *CDKN2A* were associated with poor outcome, independent of cancer type. None of 14 patients with deletions in *CDKN2A* reached a PFS ≥ 6 months (p=0.004, Fisher exact test), with a median PFS of 1.7 months (95% C.I. 1.6 – 5.5). All three patients with an *MSH3* mutation experienced an objective response (p=0.0024) across different cancer types – a pancreatic acinar cell, prostate adenocarcinoma and cutaneous squamous cell carcinoma. Significant correlations were noted between best overall response and mutations in *CDK12* (p=0.019) and *NF1* (p=0.036). Mutations in *PBRM1* and *ARID1A* (members of the SWI/SNF complex) were also associated with objective response (p=0.02; Appendix 4, Supplementary Table 4) and PFS6 (p=0.036). One individual with *CD274* (PD-L1) and *PDCD1LG2* (PD-L2) co-amplification did not respond to immunotherapy (Figure 3).

Using flow cytometry, we assayed blood-based correlates of clinical outcomes. Eighty-three patients (74%) had paired evaluable PBMC samples from baseline and week 4 following treatment. Supplementary Table 6 details the proportions of immune cell types, and subsets at both timepoints by clinical outcomes. At baseline, the most robust associations with longer OS and PFS were a higher proportion of PD-1 positive CD4^+^ T-cells, and a subset PD-1^+^ CD4^+^ cTfh cells. At week 4, a robust association with better OS and PFS was seen amongst patients with a lower proportion of granulocytic myeloid-derived suppressor cells (G-MDSC) and proliferating Ki67+ double positive T-cells (Figure 5 and Appendix 5, Supplementary Table 5). From week 0 to 4, treatment upregulated markers PD-1, HLA-DR, CD38, and Ki67 in T-cells, B-cells, and NK cells, consistent with immune cell activation and proliferation (Appendix 5, Supplementary Table 6). Overall, there was a decrease in PD-L1 expressing PBMCs, particularly amongst monocytes at 4 weeks. Compared to baseline, good responses were significantly associated with decreased G-MDSC and central memory-CD8^+^ T-cells, and increased circulating HLA-DR natural killer cells (HLA-DR^+^ NK at week 4; Appendix 5, Supplementary Figure 3 and Supplementary Table 5).

**Figure 5.**
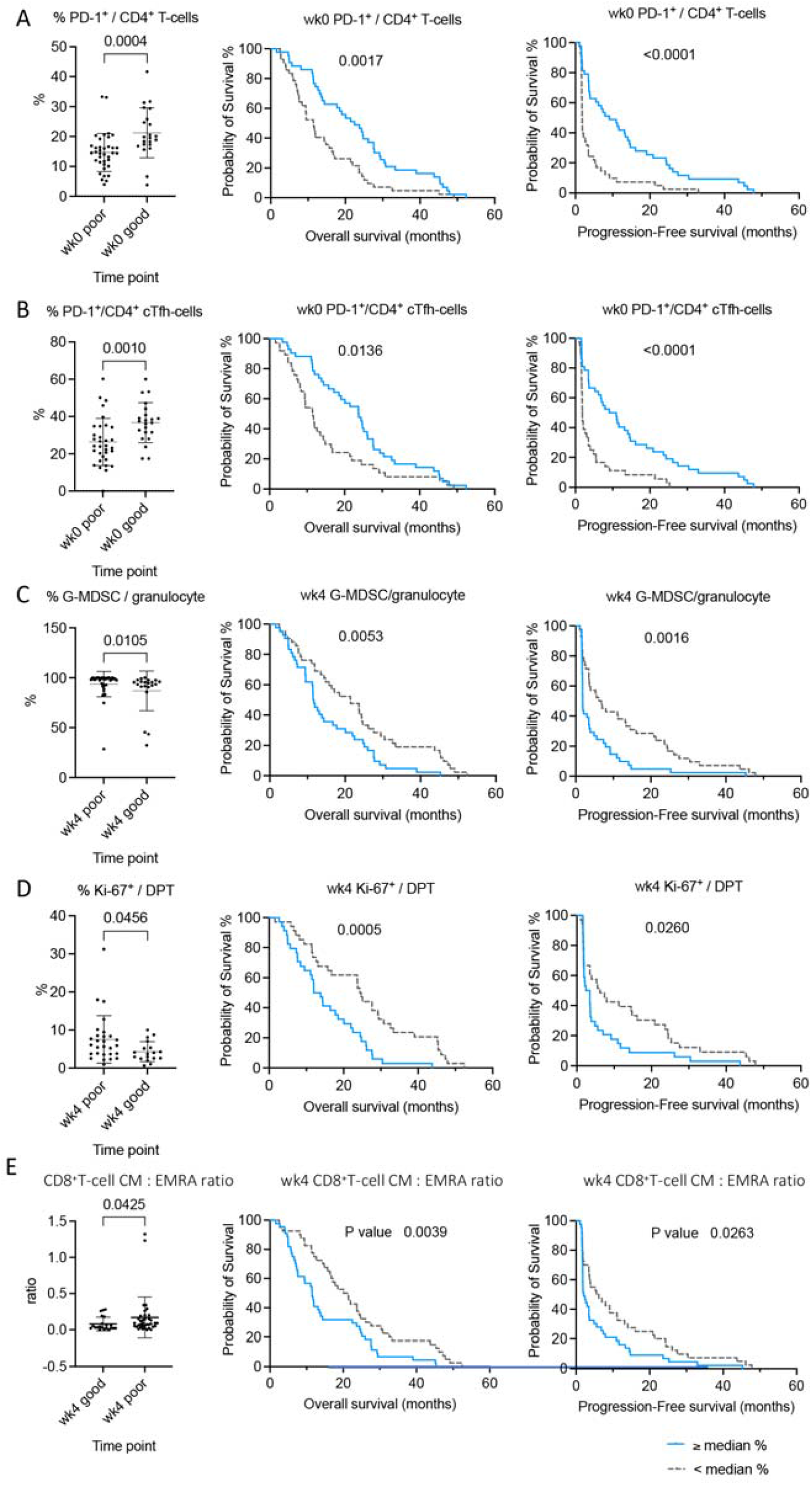
Proportionally higher PD-1^+^ CD4^+^ T-cells and PD1^+^ CD4^+^ cTfh at baseline (A&B) and lower G-MDSC Ki67+ DPT cells at week 4, and lower ratio of CD8^+^ CM: CD8^+^ EMRA (C,D and E) are associated with better outcomes. Cell subsets shown as dot plots were significantly different between poor and good, using Mann-Whitney test and Kaplan-Meier curves that associated with a significant difference in overall survival and progression-free survival of patients, using the logrank test. Time is represented in months from start date of immunotherapy.

## DISCUSSION

In this study, combination treatment with durvalumab and tremelimumab demonstrates clinical activity based on a six-month PFS rate of 32% in a tumour agnostic cohort encompassing a range of tumour histotypes. This was supported by the proportion of patients who experienced an objective response and/or a TTP2/TTP1 ratio>1.3. Objective responses occurred in 16 patients (14%) and a favourable TTP ratio in 25 patients (22%) – either a TTP ratio>1.3 where TTP1 was evaluable (16 patients), or a TTP2 ≥6 months for patients with a non-evaluable TTP1 (9 patients). Median PFS was 2.9 months and median OS was 11.9 months, with improvement in patient reported otcomes for those who remained progression-free at 6 months. Significant improvements in PFS were seen amongst patients with tumour cell PD-L1 expression ≥1% and intermediate/high TMB, while tumour cell PD-L1≥1% alone was associated with an OS advantage. Proportionally more PD1+ CD4+ T-cells at baseline and an increase in the proportion of PD-1+ CD4+ T-cells and PD-1+ cTfh following exposure to D+T significantly correlated with improved PFS and OS. Treatment was well tolerated, with no new safety concerns.

Ten patients experienced a durable objective response, including many rare histotypes, which comprised 84% of participants in this trial. To date, most trials of dual checkpoint blockade have reported on more common cancer types in populations of defined histotypes. Two studies are currently evaluating durvalumab with or without tremelimumab in advanced, refractory cancers (NCT02537418), or in combination with chemotherapy (NCT02658214) (24,25). Of note, our study had a high proportion of sarcoma patients. The Alliance A091401 study (nivolumab with or without ipiliumumab) recruited patients with metastatic, locally advanced, or unresectable sarcoma (26). Similar to our observations, 16% of participants experienced objective responses to doublet immunotherapy, including leiomyosarcoma (n=2 of 14; 14%), myxofibrosarcoma, malignant fibrous histiocytoma (n=2 of 6; 33%) and an angiosarcoma (n=1 of 3; 33%) (390). In our cohort, responses were seen in liposarcoma (n=1 of 3; 33%), leiomyosarcoma (n=1 of 10; 10%), and epithelioid hemangioendothelioma (n=2 of 2; 100%). The PFS and OS for our sarcoma patients (n=46) of 2.92 and 13.11 months was comparable to the PFS and OS of 4.1 months and 14.3 months reported in this trial (23). In general, response rates were lower than for cancers where doublet immunotherapy is considered standard of care in the first line setting. In advanced melanoma, the objective response rate with ipilumumab and nivolumab was 58%, median PFS 11.5 months and OS of 36.9 months (27-29). Similarly, amongst patients with intermediate/ high risk renal cell carcinoma, ipilumumab and nivolumab yielded an objective response rate of 42%, median PFS of 11.2 months and OS of 48.1 (27). Whether these differences are due to the late line of therapy in the current trial, to the drugs used, or the underlying immune responsiveness, is not clear.

The dose and schedule of Durvalumab (D) and Tremelimumab (T) used in this study (T75+D) was tested in a phase 1/2 trial (NCT02519348) and compared to a different dose of Tremelimumab (T300+D) in patients with advanced hepatocellular carcinomas (30). The primary study endpoint was safety and this was deemed acceptable based on ≥ grade 3 toxicity rates between 21 and 44% across these regimens, comparable with the rates seen in our study. The objective response rate observed with the combination of D+T was 24% for T300+D and 10% for T75+D, while the median OS was 19 months and 11 months, respectively. This suggests dose and scheduling of the combination of D+T may be important, at least in some tumour types. The randomised phase 3, HIMALAYA trial in unresectable hepatocellular carcinomas (NCT03298451) has just reported superior efficacy for T300+D compared with sorafenib (31).

Given the histological and genomic diversity of the study cohort, pre-specified exploratory biomarkers aimed to identify unifying correlates of clinical benefit. While we attempted to enrich our study cohort for tumours with intermediate/high TMB, ultimately only 19 patients with an intermediate/high TMB were treated on the trial. This was due to the use of multiple sequencing assays for screening, a lack of standardised bioinformatic pipelines for filtration of variants and cut-offs by assay, and evolving harmonisation strategies to compare TMB across assays. Despite these challenges, the small number of patients with a TMB ≥10 demonstrated significantly improved PFS (29.1 versus 2.0 months) and OS (17.2 versus 11.5 months), consistent with previous studies suggesting that TMB is a tumour agnostic biomarker predicting response to immunotherapy. Those patients with poor OS despite high TMB either had histotypes where there appears to be no relationship between CD8 T-cell levels and neoantigen load (glioblastomas) (32) or carcinomas where TMB in the absence of mismatch repair deficiency remains a controversial biomarker (intestinal adenocarcinomas) (33). Notably, no patients with high grade gliomas responded in this study. High tumour PD-L1 expression was also associated with significant improvements in PFS and OS across cancer histotypes, while TILs was not. CGP also revealed novel insights with both prognostic and/ or predictive potential. CDKN2A deletions demonstrated poor prognosis across several cancer types, in line with the association of CDKN2A deletions in IDH mutant gliomas (34). The association of MSH3 and objective responses is also interesting and potentially warrants inclusion into standard IHC testing.

In addition to these recognised biomarkers, blood-based immune phenotyping revealed new potential predictive markers. Previous studies have suggested associations between response to immunotherapy and subsets of CD8+ T-cells and myeloid suppressor cells (35). Treatment associated reductions in myeloid suppressor cells and central memory CD8^+^ T-cells, and increased HLA-DR^+^ NK cells, were associated with response. Amongst myeoid suppressors (MDSC), granulocytic subsets in the tumour microenvironment and the peripheral blood may indicate cytotoxic CD8 T-cell activation (36). The association between response and reduced central memory (Cm) CD8^+^ cells at week 4 may indicate treatment induced differentiation into effector memory CD8^+^ cells, with increased CD8+ EMRA cells noted at week 4 (37). HLA-DR^+^ NK cells combine characteristics of NK cells and DCs, prducing proinflammatory cytokines including INFγ and TNFα (38), although their role in response to immunotherapy remains unclear. Interestingly, baseline PD-1^+^ CD4^+^ cTfh cell numbers correlated with better outcomes. In a recent syngeneic mouse study, anti-PD-L1 therapy induced a population of cTfh with enhanced B-cell activation capacity that participated in the host immune response to treatment (39). cTfh cells may play roles in chronic-phase heart transplant rejection, graft versus host disease, and systemic lupus erythematosus (38-41). Blood-based biomarkers represent a practical and dynamic measure of treatment response in real time, and these observations warrant further investigation.

A strength of this study is its histotype-agnostic nature, including a diverse range of under-studied histologies, often with limited treatment options and poor outcomes (42,43). The heterogeneity of the patient population introduces challenges in survival analysis in a non-randomised design. Within the MoST study, an OS of 16.9 months was observed for patients (n=342) receiving a molecularly matched treatment compared to 10.4 months for patients (n=948) receiving an unmatched treatment (manuscript in preparation). In the current study, this aligns with an OS of 17.2 months for patients with high TMB and 11.5 months for those without molecular enrichment. The use of TTP2/TTP1 allows participants to act as their own controls, aiding interpretation of stable disease in future uncontrolled, tumour agnostic, phase 2 trials. This may be particularly important for immunotherapies, where time-dependent endpoints may be a more sensitive measure of clinical benefit than objective response (44). Finally, this study underscores the value of correlative-science in shedding new light on biomarkers of response and immune-related adverse events.

Overall, this study demonstrates clinical activity for doublet immunotherapy in many understudied cancer populations. Clinical benefit was greatest amongst patients with tumour cell PD-L1 expression ≥1% and high TMB. Prospective application of these biomarkers within baskets of these rare cancers will be critical to understanding the histotype-specific impact of this treatment on clinical outcomes.

## Data Availability

All data produced in the present study are available upon reasonable request to the authors.

