## Appendix for "A signal-seeking Phase II trial of Durvalumab and Tremelimumab Focused on Advanced, Rare and Less Common Cancers"

**Appendix 1.**

For the CCPv2/2.2 panel, sufficient information was not available to harmonise the TMB measures. Therefore, the distribution of TMB obtained from the CCPv2/2.2 assays was examined to identify only two patients with a clearly high TMB (56.4 and 75.8 mutations/ megabase) whilst the remaining values were all clustered together closely, ranging from 0 and 5.8 mutations/megabase.

For TST170, the number of non-synonymous mutations/ megabase was calculated by normalising the WES-TMB value to 35 megabases according to method by Endris et al (10), before further conversion to the FMI-equivalent.

**Supplementary Table 1.** Harmonisation of tumour mutational burden measurements across sequencing panels

| **Substudy ID** | **Sequencing assay** | **TMB at time of MTB issue (muts/Mb)** | **TMB at time of study analysis (muts/Mb)** | **Adjusted FMI-equivalent TMB (muts/ Mb)** | **Final TMB grouping across panels** |
| --- | --- | --- | --- | --- | --- |
| T003 | CCPv2 |  | 5.8 |  | L |
| T007 | CCPv2 |  | 1.9 |  | L |
| T009 | CCPv2 |  | 56.4 |  | H |
| T010 | CCPv2 |  | 4.5 |  | L |
| T016 | CCPv2 |  | 0.6 |  | L |
| T017 | CCPv2 |  | 0.6 |  | L |
| T021 | CCPv2.2 |  | 1.3 |  | L |
| T022 | CCPv2 |  | 1.3 |  | L |
| T023 | CCPv2.2 |  | 5.1 |  | L |
| T025 | CCPv2.2 |  | 1.9 |  | L |
| T029 | CCPv2.2 |  | 4.5 |  | L |
| T032 | CCPv2.2 |  | 3.8 |  | L |
| T033 | CCPv2.2 |  | 0 |  | L |
| T034 | CCPv2.2 |  | 0.6 |  | L |
| T035 | CCPv2.2 |  | 1.3 |  | L |
| T036 | CCPv2.2 |  | 3.2 |  | L |
| T038 | CCPv2.2 |  | 0.6 |  | L |
| T039 | CCPv2 |  | 1.3 |  | L |
| T040 | CCPv2.2 |  | 1.9 |  | L |
| T041 | CCPv2 |  | 0.6 |  | L |
| T043 | CCPv2 |  | 0.6 |  | L |
| T044 | CCPv2 |  | 1.3 |  | L |
| T045 | CCPv2.2 |  | 5.7 |  | L |
| T046 | CCPv2 |  | 1.9 |  | L |
| T047 | CCPv2.2 |  | 1.9 |  | L |
| T048 | CCPv2 |  | 3.2 |  | L |

| **Substudy ID** | **Sequencing assay** | **TMB at time of MTB issue (muts/Mb)** | **TMB at time of study analysis (muts/Mb)** | **Adjusted FMI-equivalent TMB (muts/ Mb)** | **Final TMB grouping across panels** |
| --- | --- | --- | --- | --- | --- |
| T050 | CCPv2.2 |  | 4.5 |  | L |
| T052 | CCPv2 |  | 1.3 |  | L |
| T053 | CCPv2.2 |  | 0.6 |  | L |
| T054 | CCPv2.2 |  | 75.8 |  | H |
| T055 | CCPv2.2 |  | 0 |  | L |
| T056 | CCPv2 |  | 4.5 |  | L |
| T057 | CCPv2 |  | 2.6 |  | L |
| T058 | TST170 |  | 2.3 | 3.6 | L |
| T059 | CCPv2.2 |  | 2.5 |  | L |
| T060 | CCPv2.2 |  | 0.6 |  | L |
| T062 | CCPv2.2 |  | 0 |  | L |
| T064 | TST170 |  | 2.3 | 3.6 | L |
| T065 | CCPv2.2 |  | 1.3 |  | L |
| T067 | TST170 | 11.3 | 0 | 0.9 | L |
| T068 | TST170 | 2.3 | 0 | 0.9 | L |
| T070 | TST170 | 15.8 | 4.5 | 8.7 | L |
| T071 | TST170 | 38.3 | 9 | 19.2 | H |
| T073 | TST170 | 13.5 | 4.5 | 8.7 | L |
| T074 | TST170 | 11.3 | 4.5 | 8.7 | L |
| T075 | TST170 | 4.5 | 0 | 0.9 | L |
| T076 | TST170 | 11.3 | 4.5 | 8.7 | L |
| T077 | TST170 | 11.3 | 0 | 0.9 | L |
| T078 | TST170 | 11.3 | 6.8 | 14.1 | H |
| T079 | TST170 | 11.3 | 0 | 0.9 | L |
| T080 | TST170 | 11.3 | 4.5 | 8.7 | L |
| T081 | TST170 | 15.8 | 2.3 | 3.6 | L |
| T082 | TST170 | 18 | 4.5 | 8.7 | L |
| T083 | TST170 | 18 | 2.3 | 3.6 | L |
| T084 | TST170 | 15.8 | 2.3 | 3.6 | L |
| T086 | TST170 | 22.5 | 4.5 | 8.7 | L |
| T087 | TST170 | 13.5 | 2.3 | 3.6 | L |
| T088 | TST170 | 36 | 6.8 | 14.1 | H |
| T090 | FMI | Intermediate | 9 | 9.0 | L |
| T091 | CCPv2.2 | 12.1 | 4.5 |  | L |
| T092 | FMI | High | 59 | 59.0 | H |
| T093 | TST170 | 13.5 | 2.3 | 3.6 | L |
| T095 | FMI | High | 204 | 204.0 | H |
| T096 | FMI | Intermediate | 6 | 6.0 | L |
| T097 | TST170 | 6.8 | 11.3 | 24.6 | H |
| T098 | FMI | Intermediate | 9 | 9.0 | L |

| **Substudy ID** | **Sequencing assay** | **TMB at time of MTB issue (muts/Mb)** | **TMB at time of study analysis (muts/Mb)** | **Adjusted FMI-equivalent TMB (muts/ Mb)** | **Final TMB grouping across panels** |
| --- | --- | --- | --- | --- | --- |
| T099 | TST170 | 36 | 13.5 | 29.7 | H |
| T100 | TST170 | 20.3 | 4.5 | 8.7 | L |
| T101 | TST170 | 18 | 4.5 | 8.7 | L |
| T102 | TST170 | High | 11.3 | 24.6 | H |
| T103 | TST170 | Intermediate | 6.8 | 14.1 | H |
| T104 | TST170 | 20.3 | 15.8 | 35.1 | H |
| T105 | FMI | TMB low CD274 (PD-L1) amp | 5 | 5.0 | L |
| T106 | FMI | High | 23 | 23.0 | H |
| T107 | TST170 | Intermediate | 6.8 | 14.1 | H |
| T108 | TST170 | High | 2.3 | 3.6 | L |
| T109 | TST170 | High | 0 | 0.9 | L |
| T110 | TST170 | High | 13.5 | 29.7 | H |
| T111 | FMI | High | 23 | 23.0 | H |
| T112 | FMI | Intermediate | 115 | 115.0 | H |
| T113 | FMI | Intermediate | 15 | 15.0 | H |
| T114 | TST170 | 87.9 | 69.8 | 161.1 | H |
| Shaded cells indicate unavailable data. **Abbreviations:** Muts/Mb – mutations per megabase. | | | | | |

**Appendix 2 Prior treatments used for time to progression (TTP1) calculations**

**Supplementary Table 2.** TTP1 and calculated TTP2/1 ratio

| **Subject ID** | **Previous treatment** | **Previous end reason** | **TTP1 (days)** | **TTP2/TTP1 ratio** |
| --- | --- | --- | --- | --- |
| T001 | Pazopanib | Adverse event | NE | NE |
| T002 | Pazopanib | Disease progression | 15 | 3.07 |
| T003 | Pazopanib | Disease progression | 167 | 1.25 |
| T004 | Dacarbazine | Disease progression | 45 | 3.78 |
| T006 | Doxorubicin + Cisplatin | Adverse event | NE | NE |
| T007 | Imatinib | Adverse event | NE | NE |
| T008 | N/A | N/A | NE | NE |
| T009 | Carboplatin + Paclitaxel | Completed scheduled treatment | 491 | 0.69 |
| T010 | Pazopanib | Disease progression | 510 | 0.17 |
| T011 | Zoladex + Bicalutamide | Disease progression | 218 | 0.22 |
| T012 | Letrozole | Patient preference | NE | NE |
| T013 | Palbociclib | Disease progression | 111 | 2.15 |
| T014 | VCD/IE (Vincristine + Doxorubicin + Cyclophosphamide + Ifosfamide + Etoposide) | Completed scheduled treatment | NE | NE |
| T015 | N/A | N/A | NE | NE |
| T016 | Carboplatin + Caelyx | Disease progression | 247 | 0.45 |
| T017 | VCD/IE | Completed scheduled treatment | ND | ND |
| T018 | Palbociclib | Disease progression | 49 | 1.06 |
| T019 | Dacarbazine | Disease progression | 42 | 3.55 |
| T020 | Gemcitabine/ Abraxane | Disease progression | 343 | 0.10 |
| T021 | HD Ifosfamide | Disease progression | NE | NE |
| T022 | Vinorelbine + Cyclophosphamide + Sirolimus | Disease progression | NE | NE |
| T023 | Paclitaxel | Disease progression | 99 | 1.07 |
| T024 | Capecitabine | Disease progression | 59 | 1.80 |
| T025 | Xilonix clinical trial | Disease progression | 62 | 5.77 |
| T026 | N/A | N/A | NE | NE |
| T027 | N/A | N/A | NE | NE |
| T028 | Medroxyprogesterone | Disease progression | 32 | 3.34 |
| T029 | Pazopanib | Disease progression | 274 | 0.62 |
| T030 | Astralagus + Ligustrum | Disease progression | 86 | 4.10 |
| T031 | Cisplatin + Gemcitabine | Completed scheduled treatment | 428 | 0.13 |
| T032 | Folfox | Disease progression | 153 | 2.33 |
| T033 | N/A | N/A | NE | NE |
| T034 | Procarbazine + Lomustine | Completed scheduled treatment | NE | NE |
| T035 | Carboplatin + Paclitaxel | Completed scheduled treatment | NE | NE |
| T036 | Temozolomide + ABT-414 | Disease progression | NE | NE |
| T037 | Carboplatin + Paclitaxel | Adverse event | NE | NE |
| T038 | Carboplatin + Paclitaxel | Disease progression | 273 | 0.20 |
| T039 | Topotecan | Disease progression | 22 | 2.45 |
| T040 | Docetaxel plus Gemcitabine | Completed scheduled treatment | 216 | 0.25 |
| T041 | Thalidomide | Adverse event | NE | NE |
| T042 | Doxorubicin | Disease progression | 28 | 5.75 |
| T043 | Tamoxifen | Disease progression | ND | ND |
| T044 | Avastin | Disease progression | 140 | 0.23 |
| T045 | Mitotane | Disease progression | 84 | 1.26 |
| T046 | N/A | N/A | NE | NE |
| T047 | Carboplatin + Gemcitabine | Disease progression | NE | NE |
| T048 | Liposomal doxorubicin | Disease progression | 95 | 11.81 |
| T049 | N/A | N/A | NE | NE |
| T050 | N/A | N/A | NE | NE |
| T051 | N/A | N/A | NE | NE |
| T052 | Doxorubicin | Disease progression | 84 | 2.55 |
| T053 | Cisplatin + Gemcitabine | Disease progression | 134 | 0.37 |
| T054 | Cisplatin + Capecitabine | Disease progression | 85 | 10.41 |
| T055 | Paclitaxel | Completed scheduled treatment | 109 | 0.45 |
| T056 | N/A | N/A | NE | NE |
| T057 | Tamoxifen + Provera | Disease progression | 133 | 0.39 |
| T058 | VCD/IE (Vincristine + Doxorubicin + Cyclophosphamide + Ifosfamide + Etoposide) | Clinician preference | 213 | 0.79 |
| T059 | Paclitaxel + Gemcitabine + Cisplatin | Disease progression | 114 | 0.29 |
| T060 | Cisplatin | Completed scheduled treatment | NE | NE |
| T061 | FOLFOX | Completed scheduled treatment | 92 | 1.18 |
| T062 | N/A | N/A | NE | NE |
| T063 | Vinblastine | Disease progression | 426 | 1.92 |
| T064 | VIDE | Disease progression | ND | ND |
| T065 | Palbociclib | Disease progression | 68 | 0.63 |
| T066 | Cediranib | Disease progression | 100 | 0.89 |
| T067 | Doxorubicin | Disease progression | 63 | 6.81 |
| T068 | N/A | N/A | NE | NE |
| T069 | Everolimus | Disease progression | 547 | 0.17 |
| T070 | Temozolomide | Completed scheduled treatment | 41 | 0.90 |
| T071 | Docetaxel + Simvastatin + Androgen deprivation | Disease progression | 97 | 8.47 |
| T073 | Doxorubicin + Olaratumab | Completed scheduled treatment | 127 | 0.50 |
| T074 | N/A | N/A | NE | NE |
| T075 | N/A | N/A | NE | NE |
| T076 | Procarbazine + Lomustine | Completed scheduled treatment | 461 | 0.12 |
| T077 | Everolimus | Other | 348 | 0.14 |
| T078 | Sonidegib | Disease progression | 105 | 1.62 |
| T079 | Capecitabine | Disease progression | 69 | 11.16 |
| T080 | Doxorubicin and ifosfamide | Completed scheduled treatment | NE | NE |
| T081 | FOLFIRI + Avastin | Other | 406 | 0.14 |
| T082 | FOLFIRI | Disease progression | 270 | 0.39 |
| T083 | Eribulin | Disease progression | 119 | 0.46 |
| T084 | Depatuximab + Temozolomide | Disease progression | 145 | 0.35 |
| T085 | FOLFOX | Disease progression | 110 | 0.46 |
| T086 | Capecitabine | Adverse event | ND | ND |
| T087 | FOLFIRI | Completed scheduled treatment | 127 | 1.91 |
| T088 | FLOT | Completed scheduled treatment | NE | NE |
| T089 | Liposomal Doxorubicin | Disease progression | 215 | 0.14 |
| T090 | Doxorubicin | Disease progression | NE | NE |
| T091 | Vincristine + Irinotecan + Temozolomide + Bevacizumab | Completed scheduled treatment | NE | NE |
| T092 | Lormustine (CCNU) + Procarbazine | Disease progression | 56 | 0.70 |
| T093 | N/A | N/A | NE | NE |
| T094 | Cisplatin + Gemcitabine | Completed scheduled treatment | 192 | 0.30 |
| T095 | Gemcitabine + Abraxane | Disease progression | 128 | 4.62 |
| T096 | Cisplatin + Gemcitabine | Disease progression | 50 | 5.54 |
| T097 | Doxorubicin | Completed scheduled treatment | NE | NE |
| T098 | Phase1 PARPi Trial SHR2162 | Disease progression | 306 | 0.10 |
| T099 | FOLFIRI | Disease progression | 71 | 0.72 |
| T100 | FOLIFIRI + bevacizumab | Adverse event | NE | NE |
| T101 | 5FU + folinic acid | Completed scheduled treatment | NE | NE |
| T102 | Doxorubicin + Cisplatin + Methotrexate | Completed scheduled treatment | NE | NE |
| T103 | Temozolomide | Completed scheduled treatment | NE | NE |
| T104 | Vincristine + cyclophosphamide + doxorubicin | Adverse event | NE | NE |
| T105 | Pazopanib | Disease progression | 180 | 0.28 |
| T106 | Cisplatin + Etoposide | Completed scheduled treatment | NE | NE |
| T107 | FOLFIRI | Disease progression | 344 | 0.15 |
| T108 | TAS 102 (Lonsurf) | Disease progression | 39 | 0.90 |
| T109 | FOLFOX 6 + Avastin | Disease progression | 126 | 0.38 |
| T110 | Irinotecan + Avastin | Adverse event | 102 | NE |
| T111 | N/A | N/A | NE | NE |
| T112 | Temozolomide | Disease progression | NE | NE |
| T113 | N/A | N/A | NE | NE |
| T114 | Doxorubicin | Completed scheduled treatment | NE | NE |

N/A – Not available, NE – not evaluable.

**Appendix 3 Adverse events**

**Supplementary Table 3**. A) Adjudicated serious adverse events related to study treatment B) immune-related adverse events C) Other adverse events occurring in >10% of patients

| **A. Serious adverse events** | G2 | ≥G3 |
| --- | --- | --- |
| Colitis |  | 3% |
| Pneumonitis | 1% | 2% |
| Lung infection |  | 1% |
| Pancreatitis |  | 2% |
| Hepatitis | 1% | 4% |
| Hypophysitis |  | 1% |
| Myocarditis |  | 1% |
| Infusion-related reaction | 1% |  |
| Autoimmune encephalitis |  | 1% |
| **B. Immune related adverse events** | Any grade  61 (54%) | ≥G3 |
| Rash maculo-papular | 40 (36%) | 1 (1%) |
| Rash acneiform | 4 (4%) |  |
| Pruritus | 37 (33%) | 1 (1%) |
| Hyperthyroidism | 18 (16%) |  |
| Hypothyroidism | 16 (14%) |  |
| Hyperglycaemia | 3 (3%) | 1 (1%) |
| Lipase increased | 12 (11%) | 3 (3%) |
| Amylase increased | 10 (9%) |  |
| Colitis | 4 (4%) | 3(3%) |
| Sarcoidosis | 2 (2%) |  |
| Hepatitis | 3 (3%) | 2 (2%) |
| Autoimmune encephalitis | 1 (1%) | 1 (1%) |
| Hypophysitis | 1 (1%) | 1 (1%) |
| Pneumonitis | 6 (5%) | 2 (2%) |
| Myocarditis | 1 (1%) | 1(1%) |
| Pancreatitis | 1 (1%) | 1 (1%) |
| **C. Other adverse events (≥ 10%)** | | |
| Fatigue | 80 (71%) | 2 (2%) |
| Anaemia | 34 (30%) | 1 (1%) |
| Nausea | 40 (35%) | 1 (1%) |
| Diarrhoea | 40 (35%) | 1 (1%) |
| ALT increased | 30 (27%) | 1 (1%) |
| AST increased | 25 (22%) |  |
| Pain | 15 (13%) |  |
| Cough | 15 (13%) | 2 (2%) |
| Dyspnoea | 14 (13%) | 2 (2%) |
| Anorexia | 33 (29%) |  |

**Appendix 4 Translational correlates and their association with clinical outomces**

**Supplementary Figure 1.** Area under the receiver operator characteristics analysis of PD-L1 for predicting best response and

6-month progression-free survival

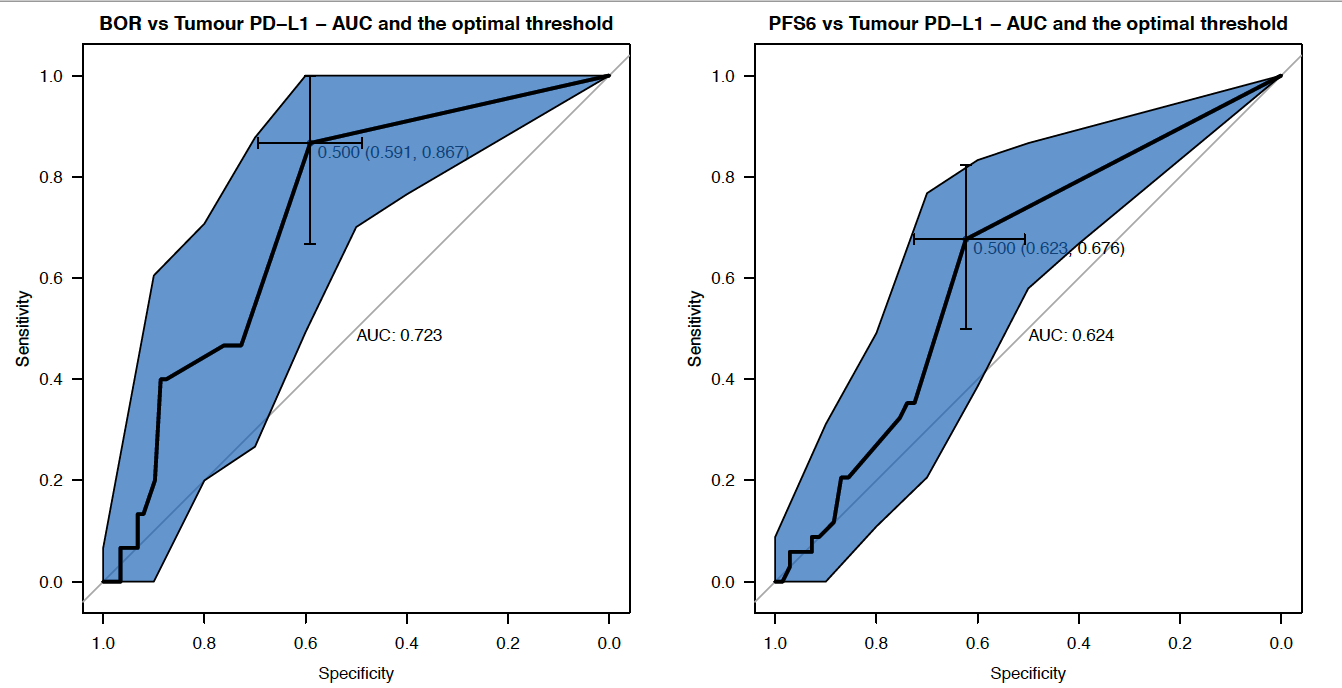

**Supplementary Figure 2.** Area under the receiver operator characteristics analysis of TMB for predicting best response and

6- month progression-free survival

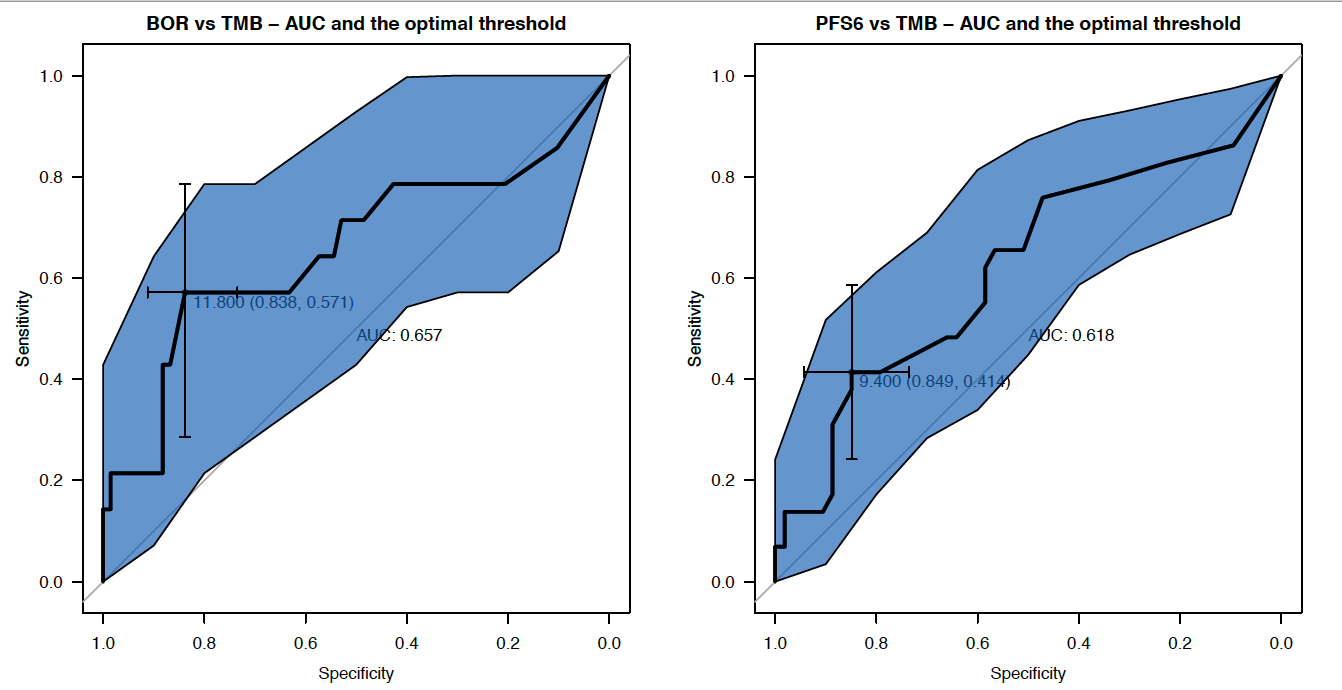

**Supplementary Table 4.** Association between best overall response and progression free survival at 6 months by alterations in specific gene and gene groups.

|  | Best overall response | | | | | | | |  | PFS at 6 months | | | | | | | |
| --- | --- | --- | --- | --- | --- | --- | --- | --- | --- | --- | --- | --- | --- | --- | --- | --- | --- |
| Biomarker | Responder  (N=15) | | Non-responder  (N=91) | | OR | 95 CI | P | Sig |  | ≥ 6 months  (N=34) | | <6 months  (N=72) | | OR | 95 CI | P | Sig |
|  | N | (%) | N | (%) |  |  |  |  |  | N | (%) | N | (%) |  |  |  |  |
| By groups |  |  |  |  |  |  |  |  |  |  |  |  |  |  |  |  |  |
| MMR genes | 3 | (23%) | 10 | (77%) | 2.02 | (0.49–8.42) | 0.39 |  |  | 6 | (46%) | 7 | (54%) | 1.99 | (0.61–6.46) | 0.34 |  |
| *ARID1A* lof | 2 | (25%) | 6 | (75%) | 2.18 | (0.4–11.97) | 0.32 |  |  | 4 | (50%) | 4 | (50%) | 2.27 | (0.53–9.67) | 0.27 |  |
| *CDKN2A* del | 0 | (0%) | 14 | (100%) |  |  | 0.21 |  |  | 0 | (0%) | 14 | (100%) |  |  | 0.0043 | ** |
| SWI/SNF lof | 3 | (60%) | 2 | (40%) | 11.1 | (1.7–73.49) | 0.02 | * |  | 4 | (80%) | 1 | (20%) | 9.47 | (1–88.26) | 0.036 | * |
| *CDK12* lof | 2 | (100%) | 0 | (0%) |  |  | 0.02 | * |  | 2 | (100%) | 0 | (0%) |  |  | 0.1 |  |
| By genes |  |  |  |  |  |  |  |  |  |  |  |  |  |  |  |  |  |
| *TP53* | 7 | (15%) | 40 | (85%) | 1.12 | (0.37–3.34) | 1 |  |  | 14 | (30%) | 33 | (70%) | 0.827 | (0.36–1.89) | 0.68 |  |
| *CDKN2A* | 3 | (18%) | 14 | (82%) | 1.38 | (0.34–5.51) | 0.71 |  |  | 3 | (18%) | 14 | (82%) | 0.401 | (0.11–1.50) | 0.26 |  |
| *APC* | 1 | (7%) | 14 | (93%) | 0.393 | (0.048–3.23) | 0.69 |  |  | 4 | (27%) | 11 | (73%) | 0.739 | (0.22–2.52) | 0.77 |  |
| *PTEN* | 2 | (15%) | 11 | (85%) | 1.12 | (0.22–5.63) | 1 |  |  | 5 | (38%) | 8 | (62%) | 1.38 | (0.42–4.58) | 0.75 |  |
| *TERT* | 0 | (0%) | 13 | (100%) |  |  | 0.21 |  |  | 5 | (38%) | 8 | (62%) | 1.38 | (0.42–4.58) | 0.75 |  |
| *KRAS* | 1 | (8%) | 11 | (92%) | 0.519 | (0.062–4.35) | 1 |  |  | 3 | (25%) | 9 | (75%) | 0.677 | (0.17–2.68) | 0.75 |  |
| *RB1* | 3 | (25%) | 9 | (75%) | 2.28 | (0.54–9.62) | 0.37 |  |  | 4 | (33%) | 8 | (67%) | 1.07 | (0.3–3.82) | 1 |  |
| *PIK3CA* | 1 | (11%) | 8 | (89%) | 0.741 | (0.086–6.39) | 1 |  |  | 3 | (33%) | 6 | (67%) | 1.06 | (0.25–4.54) | 1 |  |
| *ARID1A* | 2 | (25%) | 6 | (75%) | 2.18 | (0.4–11.97) | 0.32 |  |  | 4 | (50%) | 4 | (50%) | 2.27 | (0.53–9.67) | 0.27 |  |
| *CDK4* | 0 | (0%) | 7 | (100%) |  |  | 0.59 |  |  | 4 | (57%) | 3 | (43%) | 3.07 | (0.65–14.55) | 0.21 | * |
| *MDM2* | 0 | (0%) | 6 | (100%) |  |  | 0.59 |  |  | 3 | (50%) | 3 | (50%) | 2.23 | (0.43–11.65) | 0.38 |  |
| *NF1* | 3 | (50%) | 3 | (50%) | 7.33 | (1.3–40.55) | 0.036 | * |  | 4 | (67%) | 2 | (33%) | 4.67 | (0.81–26.87) | 0.082 |  |
| *TSC2* | 2 | (33%) | 4 | (67%) | 3.35 | (0.56–20.13) | 0.2 |  |  | 3 | (50%) | 3 | (50%) | 2.23 | (0.43–11.65) | 0.38 |  |
| *ATRX* | 1 | (20%) | 4 | (80%) | 1.55 | (0.16–14.93) | 0.54 |  |  | 1 | (20%) | 4 | (80%) | 0.515 | (0.055–4.79) | 1 |  |
| *CDK12* | 2 | (40%) | 3 | (60%) | 4.51 | (0.69–29.62) | 0.15 |  |  | 2 | (40%) | 3 | (60%) | 1.44 | (0.23–9.03) | 0.65 |  |
| *FBXW7* | 0 | (0%) | 5 | (100%) |  |  | 1 |  |  | 1 | (20%) | 4 | (80%) | 0.515 | (0.055–4.79) | 1 |  |
| *IDH1* | 1 | (20%) | 4 | (80%) | 1.55 | (0.16–14.93) | 0.54 |  |  | 1 | (20%) | 4 | (80%) | 0.515 | (0.055–4.79) | 1 |  |
| *EGFR* | 0 | (0%) | 4 | (100%) |  |  | 1 |  |  | 0 | (0%) | 4 | (100%) |  |  | 0.3 |  |
| *MSH6* | 1 | (25%) | 3 | (75%) | 2.1 | (0.2–21.59) | 0.46 |  |  | 1 | (25%) | 3 | (75%) | 0.697 | (0.07–6.96) | 1 |  |
| *PMS2* | 0 | (0%) | 4 | (100%) |  |  | 1 |  |  | 1 | (25%) | 3 | (75%) | 0.697 | (0.07–6.96) | 1 |  |
| *AKT3* | 1 | (33%) | 2 | (67%) | 3.18 | (0.27–37.42) | 0.37 |  |  | 2 | (67%) | 1 | (33%) | 4.44 | (0.39–50.73) | 0.24 |  |
| *BRAF* | 0 | (0%) | 3 | (100%) |  |  | 1 |  |  | 0 | (0%) | 3 | (100%) |  |  | 0.55 |  |
| *BRCA2* | 0 | (0%) | 3 | (100%) |  |  | 1 |  |  | 2 | (67%) | 1 | (33%) | 4.44 | (0.39–50.73) | 0.24 |  |
| *CCNE1* | 0 | (0%) | 3 | (100%) |  |  | 1 |  |  | 0 | (0%) | 3 | (100%) |  |  | 0.55 |  |
| *ERBB2* | 0 | (0%) | 3 | (100%) |  |  | 1 |  |  | 0 | (0%) | 3 | (100%) |  |  | 0.55 |  |
| *MDM4* | 1 | (33%) | 2 | (67%) | 3.18 | (0.27–37.42) | 0.37 |  |  | 3 | (100%) | 0 | (0%) |  |  | 0.031 | * |
| *MLH1* | 0 | (0%) | 3 | (100%) |  |  | 1 |  |  | 2 | (67%) | 1 | (33%) | 4.44 | (0.39–50.73) | 0.24 |  |
| *MSH2* | 2 | (67%) | 1 | (33%) | 13.8 | (1.2–163.68) | 0.052 |  |  | 2 | (67%) | 1 | (33%) | 4.44 | (0.39–50.73) | 0.24 |  |
| *MSH3* | 3 | (100%) | 0 | (0%) |  |  | 0.0024 | ** |  | 3 | (100%) | 0 | (0%) |  |  | 0.031 | * |
| *MYC* | 0 | (0%) | 3 | (100%) |  |  | 1 |  |  | 0 | (0%) | 3 | (100%) |  |  | 0.55 |  |
| *PBRM1* | 2 | (67%) | 1 | (33%) | 13.8 | (1.2–163.68) | 0.052 |  |  | 3 | (100%) | 0 | (0%) |  |  | 0.031 | * |
| *PIK3R1* | 1 | (33%) | 2 | (67%) | 3.18 | (0.27–37.42) | 0.37 |  |  | 2 | (67%) | 1 | (33%) | 4.44 | (0.39–50.73) | 0.24 |  |
| *PPP2R1A* | 1 | (33%) | 2 | (67%) | 3.18 | (0.27–37.42) | 0.37 |  |  | 2 | (67%) | 1 | (33%) | 4.44 | (0.39–50.73) | 0.24 |  |

(*) indicate statistical significance at alpha < 0.05 (**) indicate statistical significance at alpha < 0.001 (**)

**Appendix 5. Immune phenotyping** **of peripheral blood mononuclear cells**

**Supplementary methods**

Blood was collected in sodium heparin tubes and isolated PBMCs were stored frozen until analysed by flow cytometry. Peripheral blood immune cell subsets were evaluated by flow cytometry using three antibody panels. The T cell panel phenotyped T cells (including CD4^+^Tregs and circulating CD4^+^Tfh cells), B cells, NK cells, and monocytes for surface checkpoint markers, maturation or activation, and proliferation. The mixed/myeloid cell panel was used for deeper subsets of monocytes, NK cells, B cells, DCs and granulocytes. The intracellular cytokines panel analysed IFNγ, IL-2, and TNFα in T cells, B cells, and monocytes. Cryopreserved PBMCs were thawed in a 37°C water bath and washed in complete RPMI media. Intracellular cytokine staining was conducted on cells cultured with Golgi plug (1 ul/ml) with or without PMA (50 ng/ml) and ionomycin (1 ug /ml) for 4 hours at 37°C/5% CO2 and washed. A fixable viability stain and Fc block (Fc1.3216) were applied on cells, followed by labelling with surface marker antibodies. PBMCs were stained in three panels with pre-titrated amounts of mixed reagents. Cells were then fixed and, for the T cell and intracellular cytokine panels only, permeabilised, followed by labeling with intracellular marker antibodies. Beads or PBMCs were also labelled with each antibody to calculate the compensation matrix for each panel. Data were acquired with an LSR II Fortessa and FACS Diva software (BD), followed by gating analyses with FlowJo v10.6.2 (BD). Of the 112 patients enrolled, 83 had paired evaluable PBMC at baseline and 4 weeks post-treatment available for FACS analyses. Good (CR, PR, SD <12 months) 22 patients week 0, 21 week 4, Intermediate (12 months> SD>6 months, 3 months <TTP<6 months, 25 patients week 0 and 25 at week 4, and Poor Progressive disease ( TTP<3 months), 37 at both week 0 and week 4.

**Statistical Analysis**

Due to non-normality in flow cytometric data, the Mann-Whitney test, or Kruskal-Wallis tests with Dunn’s post-test, were used to make comparisons, and P- values of <0.05 were considered statistically significant. Statistical analyses were performed using Prism v9.3.1 (GraphPad). Survival analysis were done using the median values on all cell populations identified as a threshold for stratification of patients. In populations of interest, we further triaged into tertiles to determine the strength of the association. Samples were excluded from statistical analyses if the event count of the parent was <50. Wilcoxon test was used to compare changes in paired samples from week 0 to week 4.

**Supplementary Table 5**. Flow cytometry data of peripheral blood immune cell biomarkers at week 0 and week 4. Cell types or subsets that were significantly different between outcomes (good vs poor, using Mann-Whitney and survival (overall survival OS, or progression-free survival PFS, using Log-rank tests) are highlighted (higher at baseline or 4 weeks associated with good outcomes (pink), and lower numbers associated with good outcomes in blue). Mann Whitney test was conducted on evaluable samples in week 0 (22 good, 37 poor) and week 4 (21 good, 37 poor). Survival analyses were done using median values on all cell populations identified as a threshold for stratification of patients.

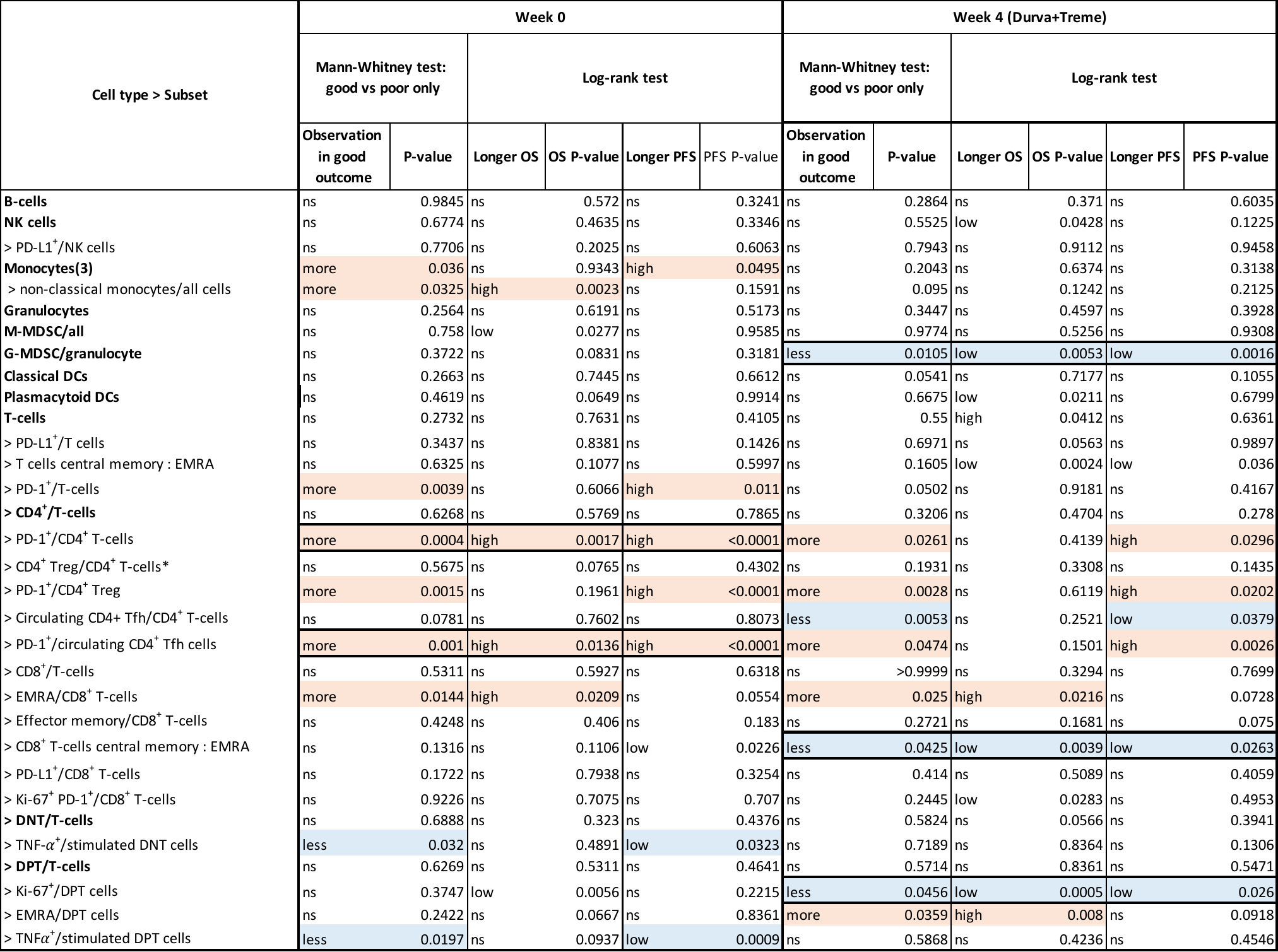

Natural killer cells (NK cells), Monocytic myeloid-derived suppressor cells (M-MDSC) Granulocytic myeloid-derived suppressor cells (G-MDSC), Dendritic cell (Dcs), Effector memory cells re-expressing CD45R (EMRA), Double negative T-cell (DNT), Double positive T-cell (DPT). Bold text denotes parental immune population. Populations are defined as percentage of all immune cells or as % parental population

**Supplementary Table 6. Effect of D+T from week 0 to week 4, whole cohort and divided into good and poor outcomes.** Flow cytometry data of peripheral blood immune cell biomarkers at week 0 and week 4, paired samples showing cell types or subsets that were significantly (highlighted in green (increased) or yellow (decreased) expression) different in response to D+T overall and in good vs intermediate and poor. Wilcoxon test.

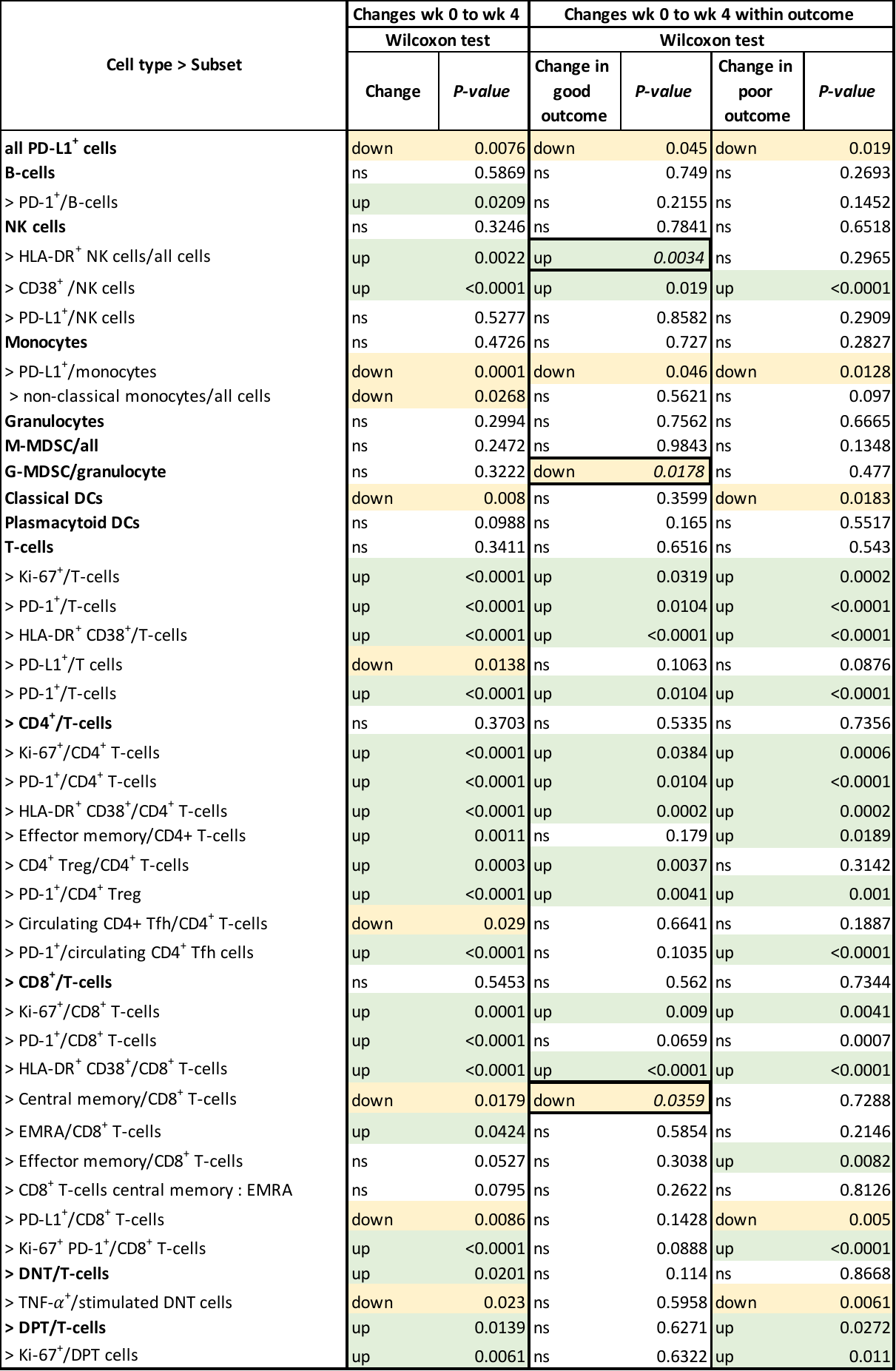

Natural killer cells (NK cells), Monocytic myeloid derived suppressor cells (M-MDSC) Granulocytic myeloid derived suppressor cells (G-MDSC), Dendritic cell (Dcs), Effector memory cells re-expressing CD45R (EMRA), Double negative T-cell (DNT), Double positive T-cell (DPT). Bold text denotes parental immune population. Populations are defined as percentage of all immune cells or as % of the parental population. Wilcoxon test used to compare paired samples from week 0 to week 4. Good (objective response, SD>12 months) 22 patients week 0, 21 week 4, Intermediate (SD, TTP> 3 month) 25 patients week 0 and 25 weeks 4, and Poor (Progressive disease, TTP<3 months) 37 at both week 0 and week 4.

**Supplementary Figure 3.** Populations where week 0 to week 4 change occurs only in good outcome patients. Flow cytometry data of peripheral blood immune cell biomarkers at week 0 and week 4, paired-samples key subsets that were significantly different at week 4 in response to D+T overall and in good and poor. Wilcoxon test. *P-values significant P<0.05.*

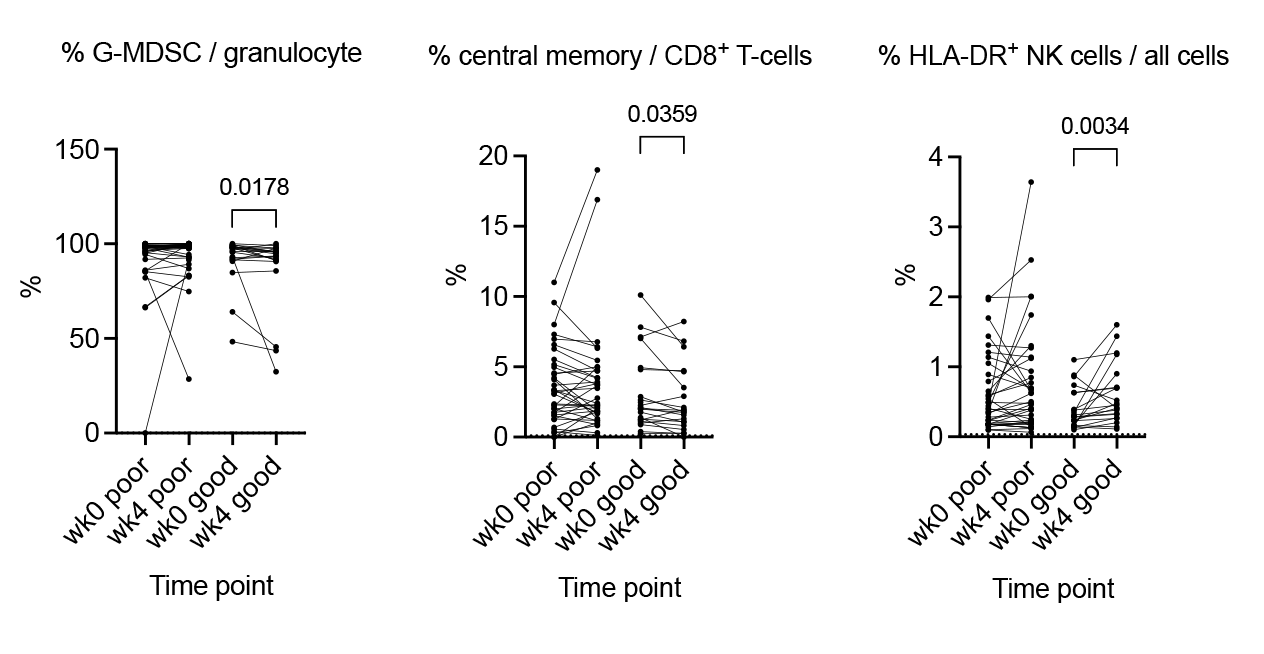

**Flow cytometry Panels**

1. T cell panel

| Marker/Reagent | Fluorophore conjugate | Clone | Source | Phenotyping |
| --- | --- | --- | --- | --- |
| Fixable Viability Stain | 700 |  | BD | - |
| Human Fc block |  | Fc1.3216 | BD | - |
| CD3 | Brilliant Violet 786 | UCHT1 | BD | Whole T cells |
| CD4 | Brilliant UV 737 | SK3 | BD | T cell subset |
| CD8a | Brilliant UV 395 | RPA-T8 | BD | T cell subset |
| CD14 | Brilliant Violet 650 | M5E2 | BD | Monocytes |
| CD19 | Brilliant Violet 711 | SJ25C1 | BD | Whole B cells |
| CD56 | PE-Cy5 | B159 | BD | NK cells, T cell activation |
| HLADR | Brilliant Violet 605 | G46-6 | BD | Activation |
| CD127 | Brilliant Violet 421 | HIL-7R-M21 | BD | T cell subset |
| CD45RA | Brilliant Blue 515 | HI100 | BD | T cell maturation |
| PDL1 | PE | MIH1 | BD | Checkpoint |
| CCR7 (CD197) | PE-CF504 | 150503 | BD | T cell maturation |
| CD25 | PE-Cy7 | M-A251 | BD | T cell activation, CD4Treg |
| CXCR5 (CD185) | AlexaFluor 647 | RF8B2 | BD | Circulating CD4Tfh |
| CD38 | APC-H7 | HB7 | BD | T cell activation |
| PD1 | Brilliant Blue 700 | EH12.1 | BD | Checkpoint, T cell activation |
| Ki67 | Brilliant Violet 486 | B56 | BD | Proliferation |

2. Mixed/Myeloid cell panel

| Marker/Reagent | Fluorophore conjugate | Clone | Source | Phenotyping |
| --- | --- | --- | --- | --- |
| Fixable Viability Stain | 700 |  | BD | - |
| Human Fc block |  | Fc1.3216 | BD | - |
| CD3 | Brilliant Violet 786 | UCHT1 | BD | Whole T cells |
| CD14 | Brilliant Violet 650 | M5E2 | BD | Monocytes |
| CD19 | Brilliant Violet 711 | SJ25C1 | BD | Whole B cells |
| CD16 | APC-H7 | 3G8 | BD | Monocyte subset, NK subset |
| CD56 | PE-Cy5 | B159 | BD | NK cells |
| CD11c | Brilliant Violet 480 | B-ly6 | BD | Classical DCs |
| HLA-DR | Brilliant Violet 605 | G46-6 | BD | Activation, antigen presentation |
| CD27 | Brilliant Violet 421 | M-T271 | BD | T cell activation, B cell maturation |
| CD10 | PE | HI10a | BD | B cell maturation |
| CD123 | PE-Cy7 | 7G3 | BD | Plasmacytoid DCs |
| CD141 | APC | 1A4 | BD | Classical DC subset |
| CD1c | Brilliant Blue 515 | F10/21A3 | BD | Classical DC subset |
| CD15 | Brilliant UV 395 | HI-98 | BD | Granulocyte |
| CD244 | PE-CF594 | 2B4 | BD | T and NK cell activation |

3. T cell / Intracellular cytokines panel

| Marker/Reagent | Fluorophore conjugate | Clone | Source | Phenotyping |
| --- | --- | --- | --- | --- |
| Fixable Viability Stain | 700 |  | BD | - |
| Human Fc block |  | Fc1.3216 | BD | - |
| CD3 | Brilliant Violet 786 | UCHT1 | BD | Whole T cells |
| CD4 | Brilliant UV 737 | SK3 | BD | T cell subset |
| CD8a | Brilliant UV 395 | RPA-T8 | BD | T cell subset |
| CD14 | Brilliant Violet 650 | M5E2 | BD | Monocytes |
| CD19 | Brilliant Violet 711 | SJ25C1 | BD | Whole B cells |
| IFNγ | APC | B27 | BD | Intracellular cytokine |
| IL2 | PE | MQ1-17H12 | BD | Intracellular cytokine |
| TNFα | FITC | MAb11 | BD | Intracellular cytokine |

4. Defining markers flow cytometry

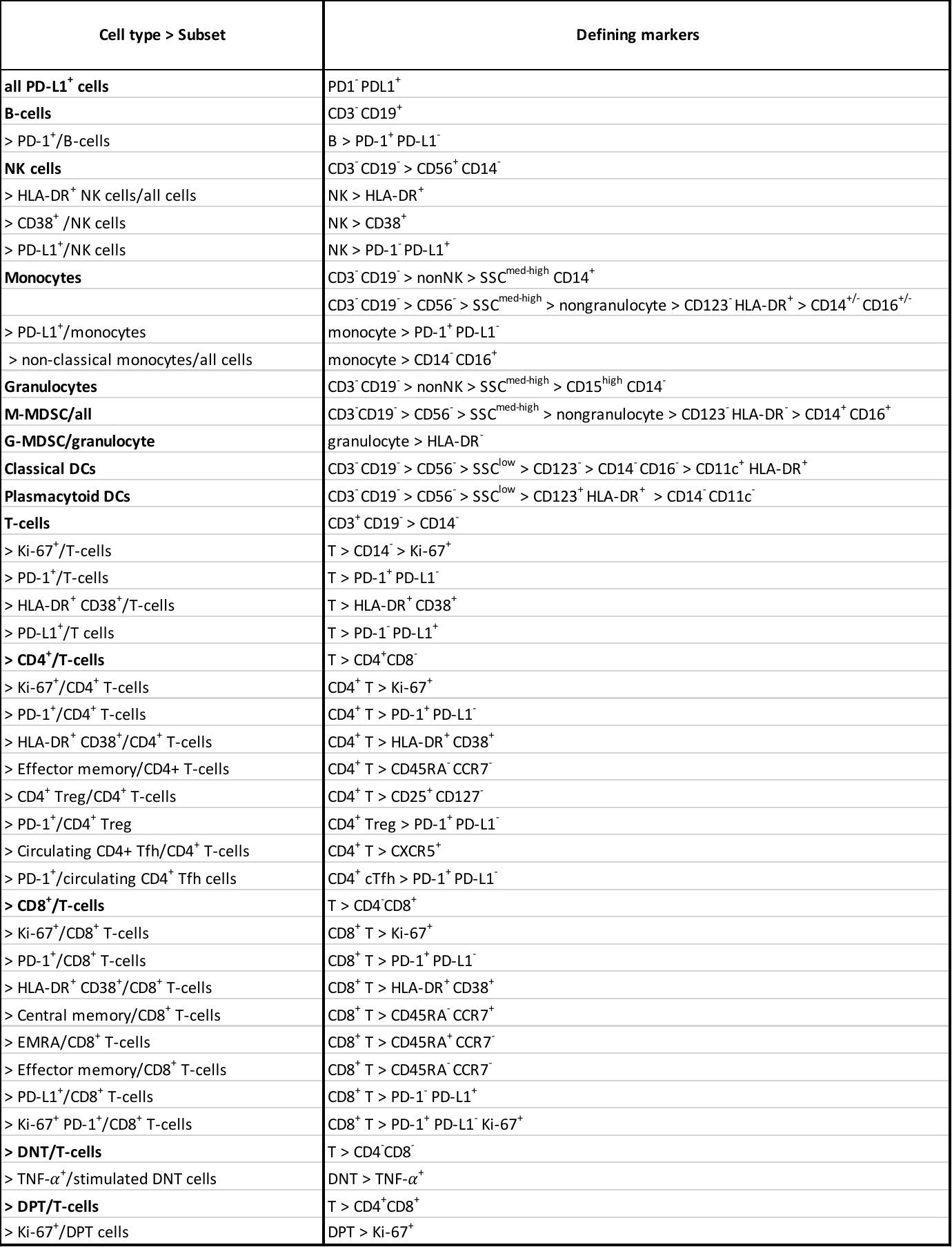

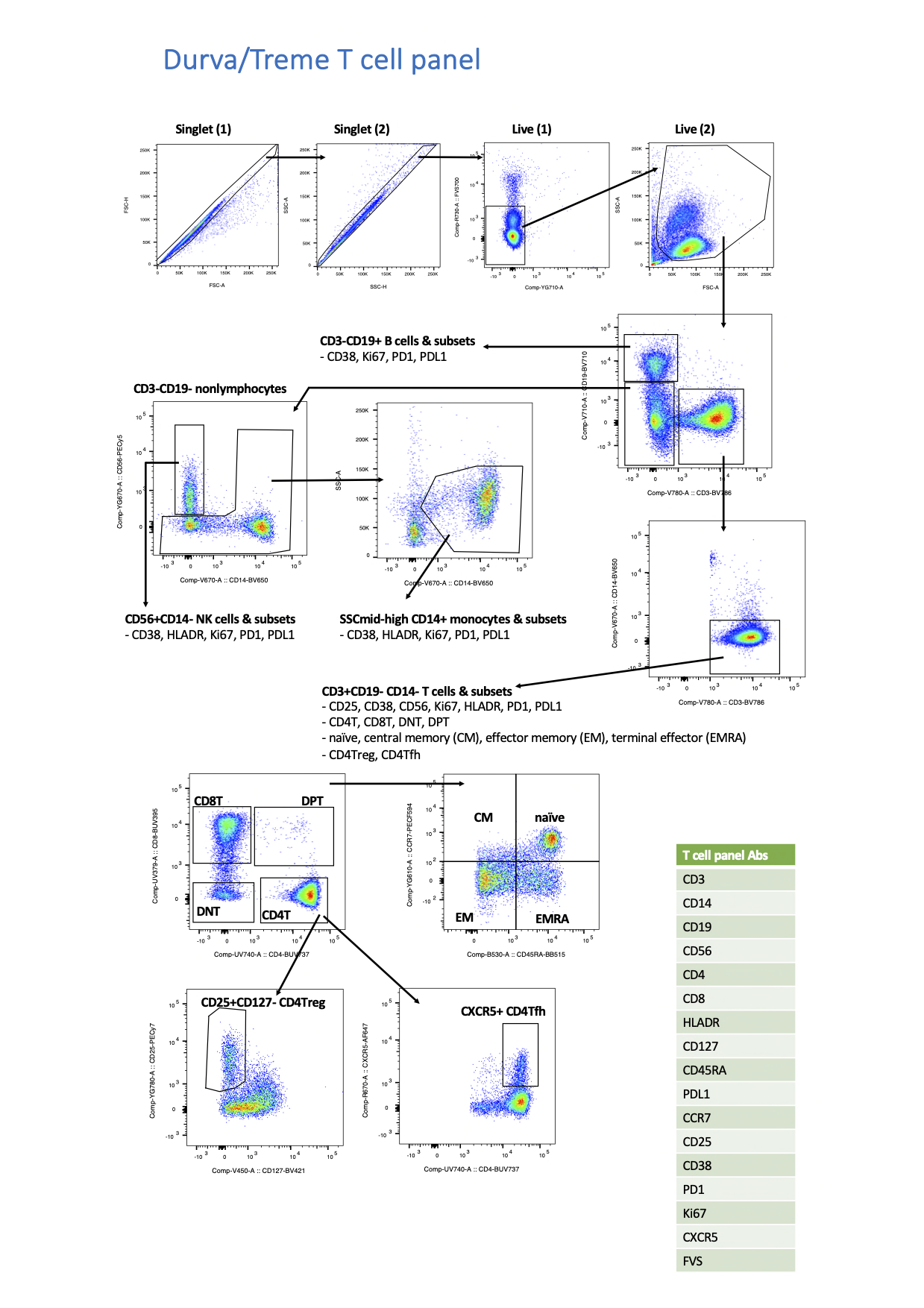
**Flow cytometry gating strategy – T-cell panel**

**Mixed myeloid panel**
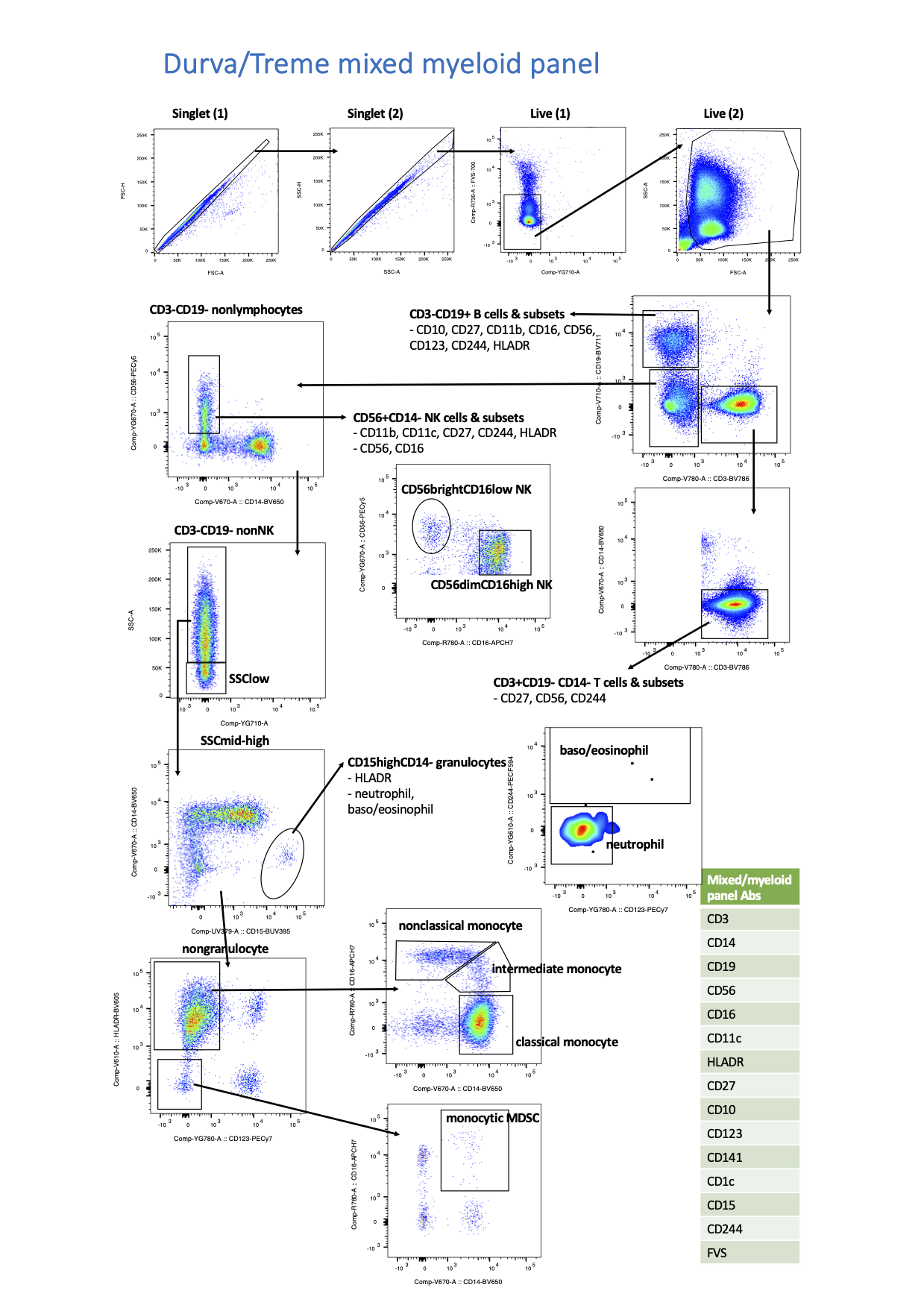

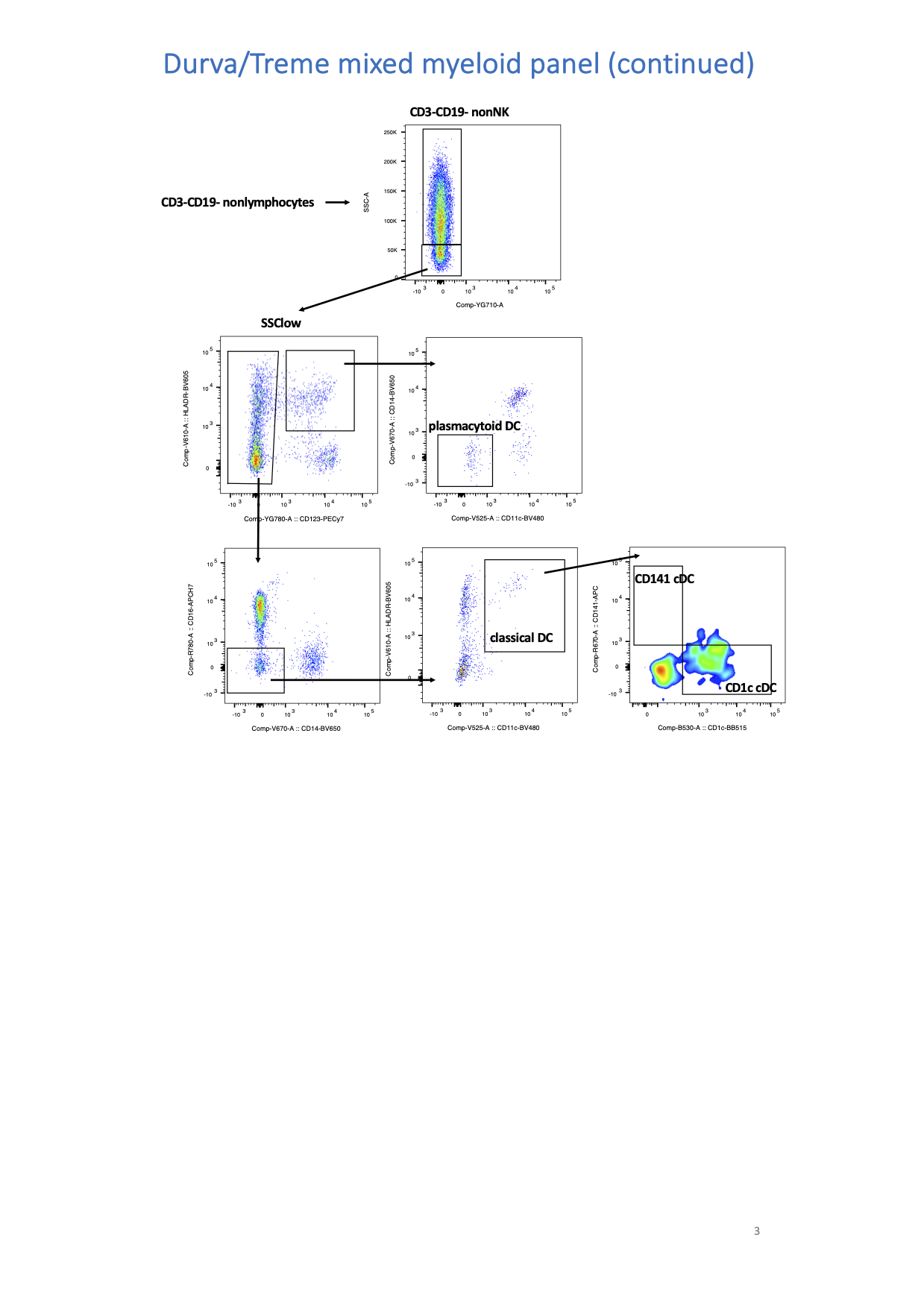
**The mixed myeloid panel continued**

**T-cell and cytokine panel**

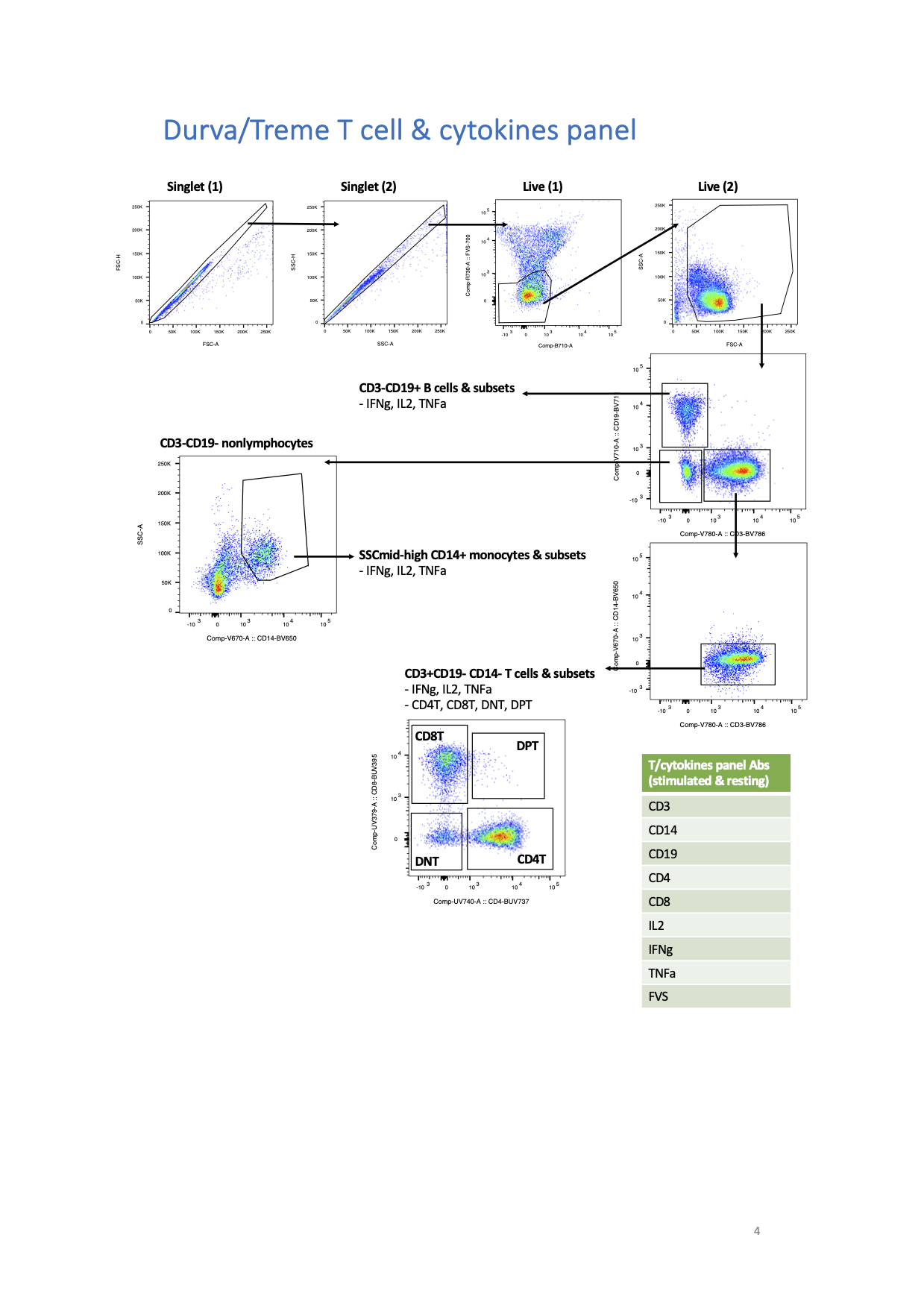
